# Functional annotation of breast cancer risk loci implicates perturbation of *FILIP1L* expression in mammary fibroblasts in influencing breast cancer risk

**DOI:** 10.64898/2026.04.09.26350488

**Authors:** Alisa Zvereva, Harriet Kemp, Andrea Gillespie, Katarzyna Tomczyk, Shirleny Romualdo Cardoso, Selin Sevgi, Kate Mackie, Vita Fedele, John Alexander, Iain Goulding, Jenny Gomm, J Louise Jones, Joseph S Baxter, Stephen J Pettitt, Christopher J Lord, Olivia Fletcher, Syed Haider, Nichola Johnson

**Affiliations:** The Breast Cancer Now Toby Robins Research Centre, The Institute of Cancer Research, London, SW3 6JB, UK; Centre for Tumour Biology, Barts Cancer Institute, Queen Mary University of London, John Vane Science Centre, Charterhouse Square, London, EC1M 6BQ, UK; The CRUK Gene Function Laboratory, The Institute of Cancer Research, London, SW3 6JB, UK

**Keywords:** breast cancer, risk locus, target gene, functional variant

## Abstract

Genome-wide association studies have led to the identification of more than 150 genomic regions that are associated with breast cancer risk. Translating these findings into a greater understanding of that risk requires identification of functional variants and target genes. Breast cancer progression and metastasis does not depend solely on cancer cell-autonomous defects; the stroma, of which fibroblasts comprise a dominant component, also has a functional role. We generated promoter capture Hi-C data in primary and immortalized mammary fibroblasts and identified 28 interaction peaks involving 116 credible causal breast cancer variants and 26 target genes that were exclusive to fibroblasts. Integrating these data with H3K27ac CUT&Tag peaks identified a potentially functional variant (rs17393059) and target gene (filamin A interacting protein 1 like (*FILIP1L*)) at the 3q12.1 breast cancer risk locus. Using genome-wide functional data in breast-relevant cell types we demonstrate that perturbation of gene expression in mammary fibroblasts may impact risk of breast cancer by a cell non-autonomous mechanism.

## Introduction

Breast cancer, which affects over 2 million women worldwide annually, has a strong heritable basis. Breast cancers typically occur in the epithelial cells that line the milk ducts and lobules of the breast with two recent studies using single cell technologies suggesting that the cell of origin for estrogen receptor-negative (ER-; basal-like) and ER+ (non-basal-like) breast cancers are luminal progenitor and mature luminal cells, respectively^1,2^ It is, however, well established that cancer progression and metastasis does not depend solely on cancer cell-autonomous defects and that the stroma, of which fibroblasts comprise a dominant component, has a functional role^3^. Specifically, a role for fibroblasts in promoting a “pre-neoplastic stroma” was reported recently^4^; the authors demonstrated that differences in gene expression in fibroblasts of *BRCA1* mutation carriers, compared to non-carriers influenced the behaviour of breast epithelial cells in these two groups, *in trans*.

Genome-wide association studies (GWAS) have led to the identification of more than 150 genomic regions that are associated with breast cancer risk^5–9^. Fine-scale mapping of these regions has identified 5,117 credible causal variants (CCVs) across 128 regions with a range of one to 375 CCVs per region^10^. The vast majority of GWAS signals map to non-protein-coding regions and are thought to influence transcriptional regulation^11,12^. Decoding GWAS risk associations, therefore, requires the identification of the subset of CCVs that are functional and the targets of these functional variants (i.e. the genes or non-coding RNAs that mediate the associations observed in GWAS). CCVs can be prioritised for further investigation by aligning them with epigenetic markers that correlate with regulatory activity such as open chromatin, active histone modifications and transcription factor (TF) binding sites. CCVs can be linked to potential target genes using chromatin interaction methods or by expression quantitative trait locus analyses (eQTL; reviewed in ^13^). Access to genome wide data sets generated in relevant cell types is fundamental to these steps.

For breast cancer specifically, there is a wealth of data in breast cancer cell lines (MCF-7, T-47D), normal immortalised breast epithelial cells (MCF10A) and primary mammary epithelial cells available through the ENCODE portal (www.encodeproject.org). For mammary fibroblasts, however, data in primary cells is sparse and there is no data in cell line(s).

We have generated genome-wide data in primary human mammary fibroblasts (HMF; promoter capture Hi-C (pCHi-C), H3K27ac CUT&Tag, RNA-seq and whole genome NEB Next EM-seq) and normal immortalised human mammary fibroblasts (GS2; promoter capture Hi-C, H3K27ac CUT&Tag and RNA-seq). To maximise the power of cell type-specific analyses we also generated data in primary human mammary luminal epithelial (HMLE) cells taken from the same women as the fibroblasts, and two commonly used cell lines (MCF10A cells and T-47D) using the same protocols. We have annotated the 5,117 breast cancer CCVs reported by Fachal *et al*^10^ with these data and describe cell type-specific interactions between CCVs and target genes at three loci. In mammary fibroblasts specifically, we identify interactions at the 3q12.1-*FILIP1L* locus; we select rs17393059 as a likely functional variant and demonstrate that targeting rs17393059 using CRISPR interference (CRISPRi) perturbs expression of *FILIP1L* in immortalized mammary fibroblasts.

## Results

We generated pCHi-C libraries in HMLE, HMF, MCF10A, T-47D and GS2 cells. For the primary cells we used biological replicates (n=2) of the two cell types, each taken from two women, for the cell lines we used technical replicates (T-47D n=2, GS2 n=2, MCF10A n=3; Methods). For all the libraries we used the Dovetail Omni-C protocol and the Dovetail Pan Promoter Enrichment Panel, sequencing the libraries to achieve between 159 and 651 million paired end reads per library. This resulted in a total of 72 million (T-47D) to 141 million (HMLE) unique on target ditags for analysis (full QC metrics are presented in Supplementary Table 1). Interaction peaks (defined as combinations of captured promoter bins and uncaptured target bins that occur more frequently than expected by chance) were called using CHiCANE^14^, resulting in 28,216 (T-47D) to 56,021 (HMLE) interaction peaks per cell type; excluding bait-to-bait interaction peaks there were 26,792 (T-47D) to 53,315 (HMLE) interaction peaks for downstream analysis (Figure 1).

**Figure 1.**
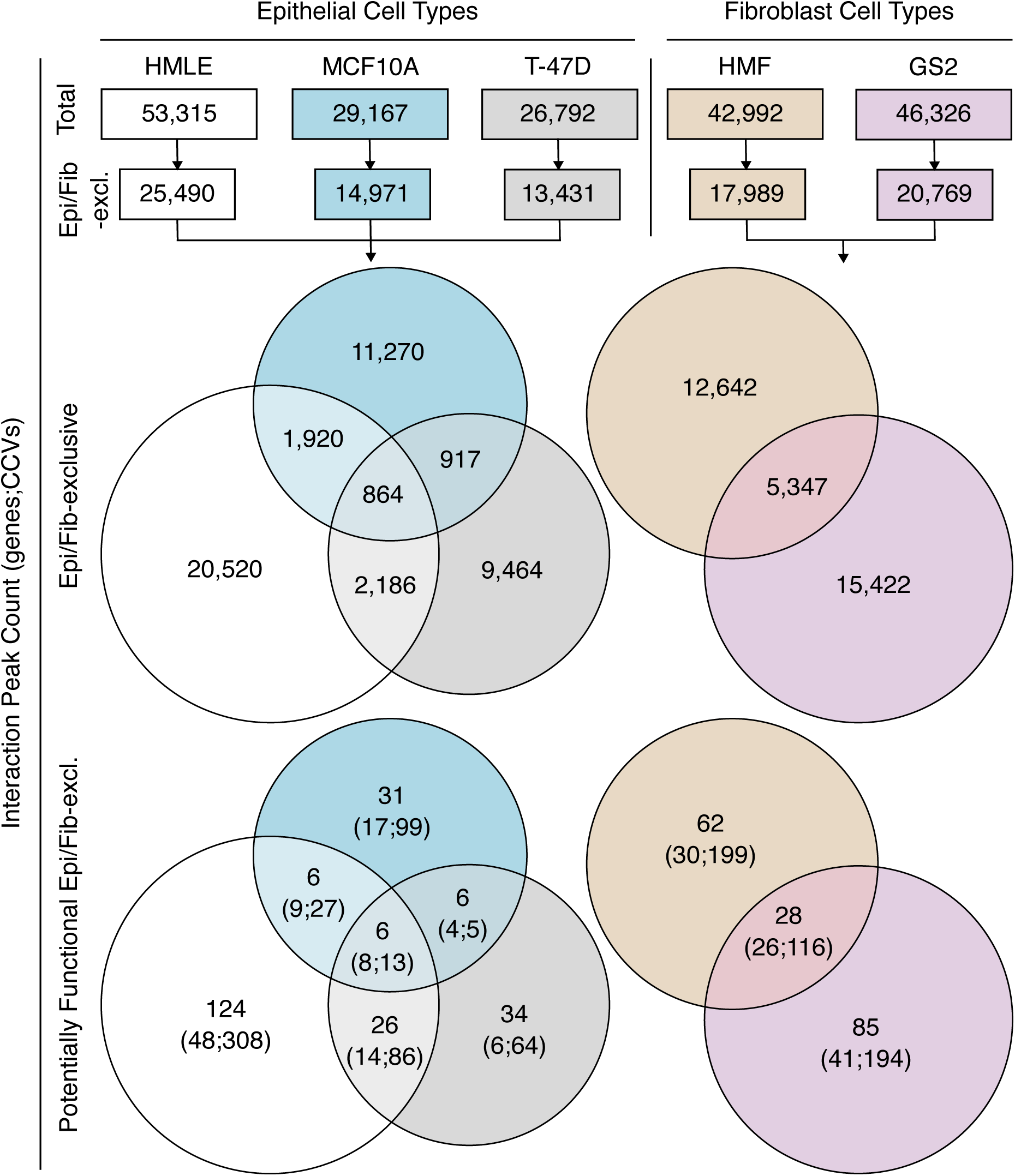
Number and cell type-specificity of interaction peaks in pCHi-C libraries from epithelial cells and fibroblasts. The total number of interaction peaks in each library (after exclusion of bait-to-bait interaction peaks; Total), the number that were exclusive to a cell type group (epithelial or fibroblast; Epi/Fib-excl.) and the number that were potentially functional (i.e. in which the captured end (mapping to a gene promoter) interacted with an uncaptured bin which colocalized with at least one credible causal variant (CCV)) are indicated.

### pCHi-C identifies potentially functional cell type-specific interaction peaks

We hypothesised that non-coding causal (as opposed to correlated) variants at breast cancer GWAS signals impact breast cancer risk by perturbing the activity of a cell type-specific enhancer and hence, expression of a target gene. Specifically, we hypothesised that a functional variant could alter binding of a transcription factor at the enhancer thereby increasing (or decreasing) expression of a target gene that has been brought into proximity with the regulatory element via “looping” ^15^. First, we used hierarchical clustering to assess the cell type-specificity of pCHi-C interaction peaks and to assess the extent to which our cell lines were good models for HMLE and HMF in the “at risk” breast (Supplementary Figure 1). GS2 clustered with HMF, supporting pCHi-C interaction peaks as showing cell type-specificity and GS2 as a reasonable model for primary mammary fibroblasts. Within the epithelial cells, however, T-47D clustered with HMLE rather than MCF10A, suggesting that the luminal epithelial breast cancer cells might be a better model for primary HMLE than “normal” immortalized mammary epithelial cells such as MCF10A that express both luminal and basal markers^16,17^.

We next selected all interaction peaks that were present in one or more of the epithelial cells but not in the fibroblasts, and vice versa. There were 25,490 (HMLE), 14,971 (MCF10A) and 13,431 (T-47D) of these cell type-specific interaction peaks in epithelial cell types. In HMF and GS2 there were 17,989 and 20,769, respectively (Figure 1). From these, we defined potentially functional interaction peaks as the subset in which the uncaptured end (i.e. the end that potentially acts as an enhancer element) contained at least one of the 5,117 CCVs reported by the Breast Cancer Association Consortium (BCAC)^10^. There were 162, 49 and 72 cell type-specific potentially functional interaction peaks involving 79, 38 and 32 different genes and 434, 144 and 168 CCVs in HMLE, MCF10A and T-47D, respectively (Figure 1, Supplementary Table 2). In HMF and GS2 there were 90 and 113 cell type-specific potentially functional interaction peaks involving 56 and 67 different genes and 315 and 310 CCVs, respectively (Figure 1, Supplementary Table 2).

To further enrich for variants that might perturb the activity of a cell type-specific enhancer, we aligned the CCVs from these interaction peaks with an epigenetic mark that correlates with active enhancer elements (H3K27ac) profiled in each of the same cell types. In HMLE, 39 (9.0%) of the CCVs that were involved in potentially functional interaction peaks also colocalized with a cell type-specific H3K27ac peak; for MCF10A and T-47D these numbers were 12 (8.3%) and 16 (9.5%), respectively (Supplementary Table 2). In HMF, 27 (8.6%) of the CCVs that were involved in potentially functional interaction peaks also colocalized with an H3K27ac peak; for GS2 the number was 26 (8.4%; Supplementary Table 2). Comparing the proportion of CCVs that mapped to an H3K27ac peak across cell types, there was no evidence of greater enrichment in any cell type (Fisher’s exact test, *P*=0.995).

The full repertoire of interaction peaks formed by each of the 5,117 BCAC selected CCVs including which CCVs colocalize with H3K27ac peaks is given in Supplementary Table 3.

### Previously reported target genes form epithelial cell type-specific interaction peaks

To determine the extent to which our data support previous functional studies we compared target genes in our data with those reported in 17 single locus studies reviewed in Romualdo Cardoso *et al*^13^. Consistent with the original analyses, we observed epithelial cell type-specific interaction peaks with *MAP3K1* at 5q11.2 in HMLE and T-47D^18^, *KLF4* at 9q31.2 in T-47D^19^, *MYC* and *PVT1* at 8q24.21 in HMLE and MCF10A^20–23^, *GATA3* at 10p14 in HMLE and T-47D^24^ and *TBX3* at 12q24.21 in HMLE^24^.

Overall, there were 38 epithelial cell type-specific interaction peaks in HMLE that were replicated in MCF10A and/or T-47D (Figure 1). From these we selected two previously uncharacterised loci at which our data implicate a putative target gene and at least one potentially functional variant.

### *TRPS1* identified as a target gene at 8q23.3

At 8q23.3 in HMLE, there is an active interactome, comprising large numbers of interaction peaks, with a subset that are relatively short-range interactions (100kb to 350kb) and that are replicated in T-47D but not in MCF10A, HMF or GS2 (Figure 2A; zoomed in view in Supplementary Figure 2A). Fine-scale mapping at this locus identified four independent signals comprising 164 (signal-1), five (signal-2), two (signal-3) and three (signal-4) CCVs^10^. In HMLE, 99 of these CCVs (mapping to 23 pCHiC bins) form interaction peaks with the *TRPS1* promoter bin (Figure 2A pink and turquoise arcs, Supplementary Table 2). The interaction peaks between 10 of these CCVs and *TRPS1* in HMLE are replicated in T-47D and two of them (rs4876626 and rs2223054; see Supplementary Table 4 for genotypes in each cell type) colocalize with an H3K27ac peak in HMLE and T-47D (rs4876626) or just T-47D (rs2223054; Figure 2A turquoise arcs, Supplementary Table 2). To determine whether either of these CCVs mapped to an element that has been implicated in driving cancer initiation, progression or metastasis we accessed data from a recent analysis of epigenetic regulation during cancer transitions across eleven tumour types^2^. Using single-nucleus chromatin accessibility data and matched single-cell or single-nucleus RNA-seq expression data, Terekhanova *et al* constructed a pan-cancer epigenetic and transcriptomic atlas across 11 cancer types and 15 normal adjacent tissues. The cancers included “non-basal” and “basal” breast cancers, for which the closest normal cells were defined as mature luminal cells and luminal progenitor cells, respectively. They compared each cancer type to all others, and cancer cells to their closest normal cells to define tissue-specific and cancer-cell-specific differentially expressed genes (DEGs) and differentially accessible chromatin regions (DACRs).

**Figure 2.**
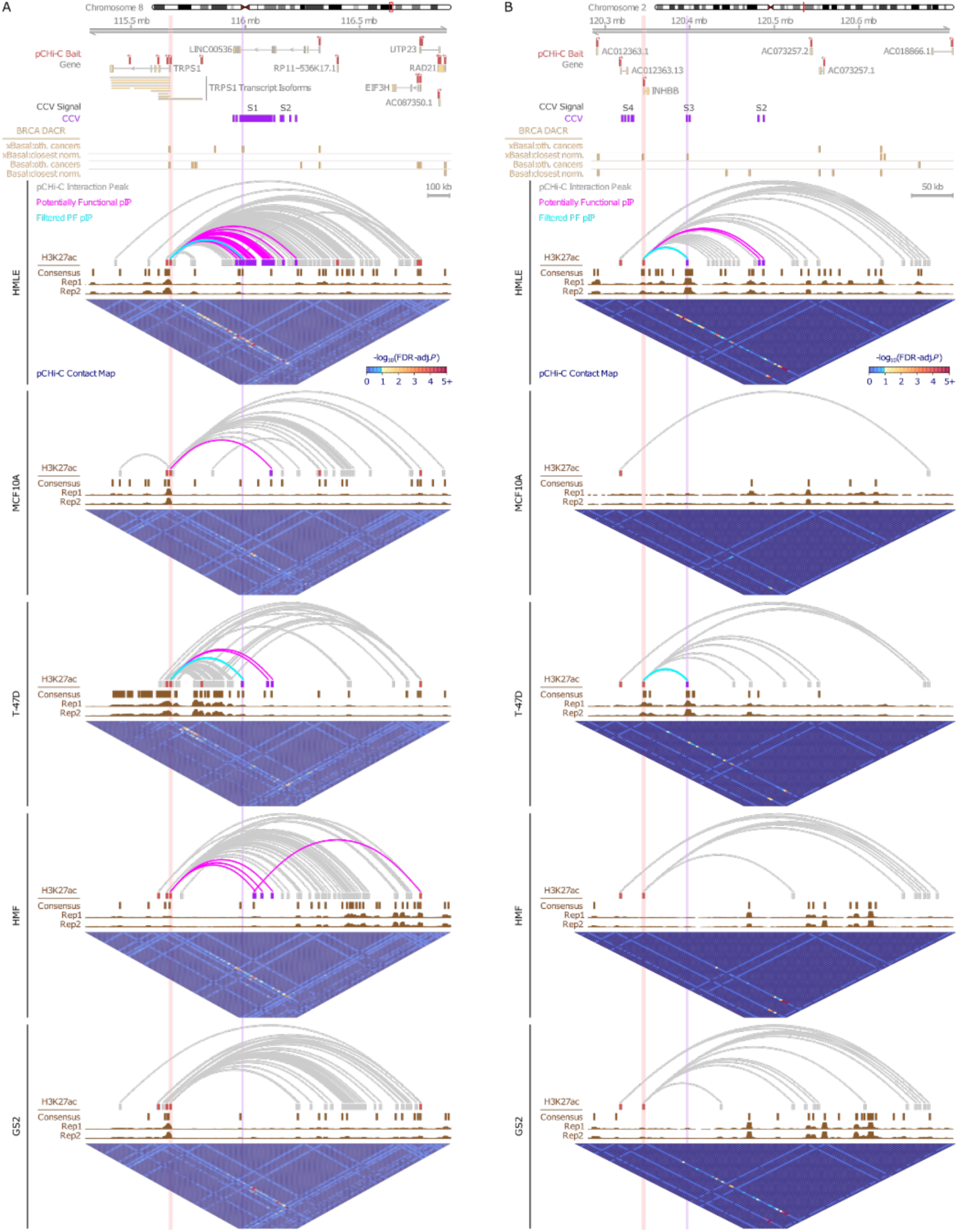
Interaction peaks at (A) 8q23.3-*TRPS1* and (B) 2q14.2-*INHBB* in epithelial cell and fibroblast pCHi-C libraries. Interaction peaks are shown as arcs (grey, pink, turquoise) and are aligned with Ensembl genes (grey/yellow; genes included on the pCHi-C panel are shown), their associated pCHi-C baits (red), and credible causal variants (CCVs, purple). H3K27ac CUT&Tag data are shown as raw reads (Rep1 and Rep2; y-axis fixed across all samples) and as called consensus peaks (dark brown). DACRs reported by Terekhanova *et al*^32^ (extended by +/- 500 bp) are shown in light brown. Potentially functional interaction peaks (i.e. those for which one end colocalises with a captured gene promoter and the other end colocalizes with a bin that harbours at least one CCV) are in pink. The subset of the potentially functional interaction peaks that are unique to a cell type group (Epi/Fib-excl.), and at which the CCV(s) also colocalize with an H3K27ac peak in at least one of the same cell types, are in turquoise. Vertical light pink and light purple lines annotate the promoter- and CCV-associated bins, respectively, of turquoise interaction peaks. Raw pCHi-C data are shown as contact maps with the -log_10_ (FDR-adjusted *P*-value) for each set of interacting genomic bins indicated on a coloured scale between deep blue (no interaction), light blue (non-significant interaction; adj.*P*³0.1), yellow (significant interaction peak; adj.*P*<0.1) and red (adj.*P*<0.00001). Contact map cells are equidistant while true genomic bins range from 269bp to 5kb. Bait-associated cells within contact maps are enhanced for visualisation. Bait-to-bait interactions are present in raw pCHi-C contact maps but excluded from interaction peak arcs.

Aligning all of the CCVs that formed potentially functional interaction peaks with DACRs (Supplementary Tables 2 and 3), both of the 8q23.3 variants (rs4876626 and rs2223054) colocalized with a DACR that shows increased accessibility in non-basal breast cancers compared to the other 10 cancers included in the analysis by Terekhanova *et al*^2^. In summary, these data implicate *TRPS1* as a target of the 8q23.3 GWAS locus; expression of *TRPS1* is increased in non-basal breast cancers compared to all other cancers (8-fold increase, *P*=0)^2^ and we have identified two CCVs that are attractive candidates for follow up studies in mature luminal epithelial cells and non-basal breast cancers or breast cancer cell lines.

### *INHBB* identified as a target gene at 2q14.2

At 2q14.2 there is an active interactome in HMLE, far fewer interactions in T-47D, HMF and GS2 and just one in MCF10A (Figure 2B; zoomed in view in Supplementary Figure 2B). Fine-scale mapping at this locus identified four independent signals comprising two (signal-1), 16 (signal-2), three (signal-3) and six (signal-4) CCVs^10^. In HMLE, 18 of these CCVs (mapping to three genomic bins) form interaction peaks with the *INHBB* promoter bin (Figure 2B pink and turquoise arcs, Supplementary Table 2). The interaction peak between two of these CCVs (rs4076654 and rs6542593; see Supplementary Table 4 for genotypes in each cell type) and *INHBB* in HMLE is replicated in T-47D and both CCVs also colocalize with an H3K27ac peak that is specific to HMLE and T-47D (Figure 2B turquoise arcs, Supplementary Table 2). In addition, rs6542593 colocalizes with a DACR that shows reduced accessibility in breast cancers compared to mature luminal cells (the normal cells that are closest to non-basal breast cancers)^2^. In summary, these data implicate *INHBB* as a target of the 2q14.2 GWAS locus; expression of *INHBB* is reduced in non-basal breast cancers compared to mature luminal cells (7% decrease, *P*=2.6 x 10^-24^)^2^ and rs6542593 is an attractive candidate as a functional variant that could be further explored in mature luminal epithelial cells and non-basal breast cancers or breast cancer cell lines.

### Fibroblast-specific interaction peaks for *FILIP1L* at 3q12.1

The main aim of this analysis was to determine whether fibroblasts could act through GWAS loci to influence breast cancer risk. Focusing on the 28 fibroblast-specific interaction peaks in HMF that were replicated in GS2 (Figure 1), we selected the 3q12.1 locus for further investigation. At this locus, there are interaction peaks originating from two *TBC1D23* promoter bins and three *FILIP1L* promoter bins (Figure 3A, 3B, 3C and Supplementary Table 2). For the purpose of our analysis multiple promoter bins for a single gene have been numbered in ascending order (“P#”) according to their 5’ to 3’ orientation. A single promoter bin can contain multiple transcription start sites (TSSs) for a gene if they are within close proximity. Fine-scale mapping at the 3q12.1 locus identified a single signal comprising 92 CCVs^10^, three of which, rs6807176, rs17393059 and rs6799379 (all mapping to a single genomic bin; see Supplementary Table 4 for genotypes in each cell type) formed interactions with two different *FILIP1L* promoter bins in HMF (P1 and P3; Figure 3B). These interactions were replicated in GS2 and were specific to the fibroblast cell types. rs6807176 also colocalized with an H3K27ac peak in HMF and HMLE, while rs17393059 colocalized with a peak in HMF, GS2 and HMLE (Figure 3A, 3C and Supplementary Table 2). Aligning all three CCVs with DNase I signals in HMF, adult skin fibroblasts, human mammary epithelial cells and breast cancer cell lines, rs17393059 mapped immediately adjacent to a region of DNase I sensitivity that was present in all fibroblast cell types but not in the epithelial cell types (Figure 3D).

**Figure 3.**
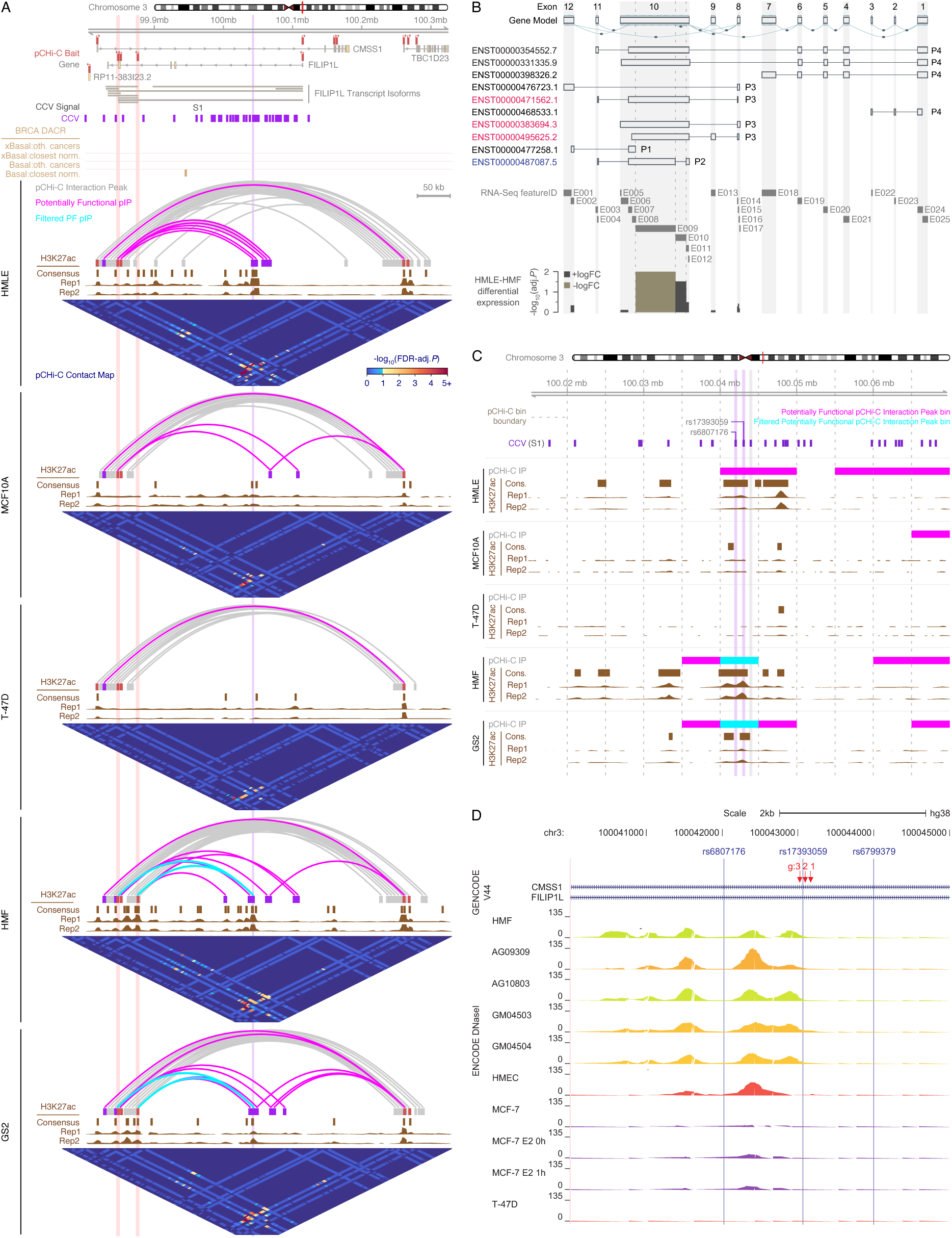
The 3q12.1-*FILIP1L* locus. (A) Interaction peaks at 3q12.1 in HMLE, MCF10A, T-47D, HMF and GS2 pCHi-C libraries are shown and are annotated as described for pCHi-C libraries in Figure 2. (B) Exon-specific expression for *FILIP1L* (ENSG00000168386.18) based on GTEx Analysis Release V8 (dbGaP Accession phs000424.v8.p2). Promoter labels based on the nomenclature used in this analysis (P1 to P4) have been added to the GTEx figure. RNA-seq features (E001 to E025) have been mapped back to the GTEx defined exons and differential expression of these features between HMLE and HMF is summarised graphically. Isoforms that are transcribed from P3 that include E010 (chr3:99,850,428-99,850,932), a 505 bp region that is present in three of the P3-transcribed isoforms as part of exon 10 in the schematic, and are translated into proteins of ∼120 kDa are in red. The isoform that is transcribed from P2 (in blue) does not include the 505 bp region (E010) and is translated into a protein of ∼90kDa. (C) Zoomed in view of the uncaptured ends of potentially functional interaction peaks are in pink and the subset that are unique to fibroblasts, and at which the CCV(s) also colocalize with an H3K27ac peak in at least one fibroblast cell type (HMF or GS2), are in turquoise. rs17393059 forms an unreplicated potentially functional interaction peak with *FILIP1L* P2 promoter in HMLE (hence the pink bar) and replicated fibroblast-specific interaction peaks with *FILIP1L* P1 and P3 promoters in HMF and GS2 (hence the turquoise bars). rs17393059 colocalizes with an H3K27ac peak in HMLE, HMF and GS2, rs6807176 colocalizes with an H3K27ac peak in HMLE and HMF. (D) Zoomed in view of the 3 CCVs (blue) that form fibroblast specific interactions with *FILIP1L*. The CCVs are aligned with DNase I signals generated in HMF, adult skin fibroblasts (AG09309, AG10803, GM04503, GM04504), human mammary epithelial cells (HMEC) and breast cancer cell lines (MCF-7, T-47D) as part of the ENCODE project (https://www.encodeproject.org/). The three sgRNAs that were used to target rs17393059 using CRISPRi are indicated (red).

We hypothesised that allelic variation at rs17393059 influences breast cancer risk by perturbing *FILIP1L* expression in fibroblasts. To further investigate this, we accessed Genotype-Tissue Expression data (GTEx; https://gtexportal.org) generated in 49 non-diseased tissue and cell types^25^; rs17393059 was associated with levels of expression of *FILIP1L* in cultured fibroblasts (NES= -0.440, *P*=1.6 x 10^-17^) but not in mammary tissue (NES=0.020, *P*=0.6; Supplementary Figure 3). It has previously been reported that *FILIP1L* expression is regulated, in part, via promoter methylation^26–31^. Comparing the proportion of methylated CpGs at *FILIP1L* promoter bins between HMLE and HMF, we found that the P3 promoter bin, which colocalizes with a CpG island, is hypomethylated in HMF compared to HMLE (Figure 4) and colocalizes with an extended region of H3K4me3 histone modification (Supplementary Figure 4). *FILIP1L* was not differentially expressed in HMLE compared to HMF overall (log_2_ fold change (FC)= -0.28, *P*=0.12) however, exon-specific analysis of RNA-seq data showed differential expression of *FILIP1L* isoforms between HMLE and HMF (Supplementary Table 5). Specifically, P3-transcribed isoforms (red) are distinguishable from P2-transcribed isoform (blue), by the presence and/or absence of E010 (chr3:99,850,428-99,850,932: Figure 3B). Thus, preferential expression of E010 containing isoforms in HMF and E009 (chr3:99,848,613-99,850,427 bps) containing isoforms in HMLE confirms a cell type-specific preference for transcription from P3 in HMF and P2 in HMLE (Supplementary Table 5).

**Figure 4.**
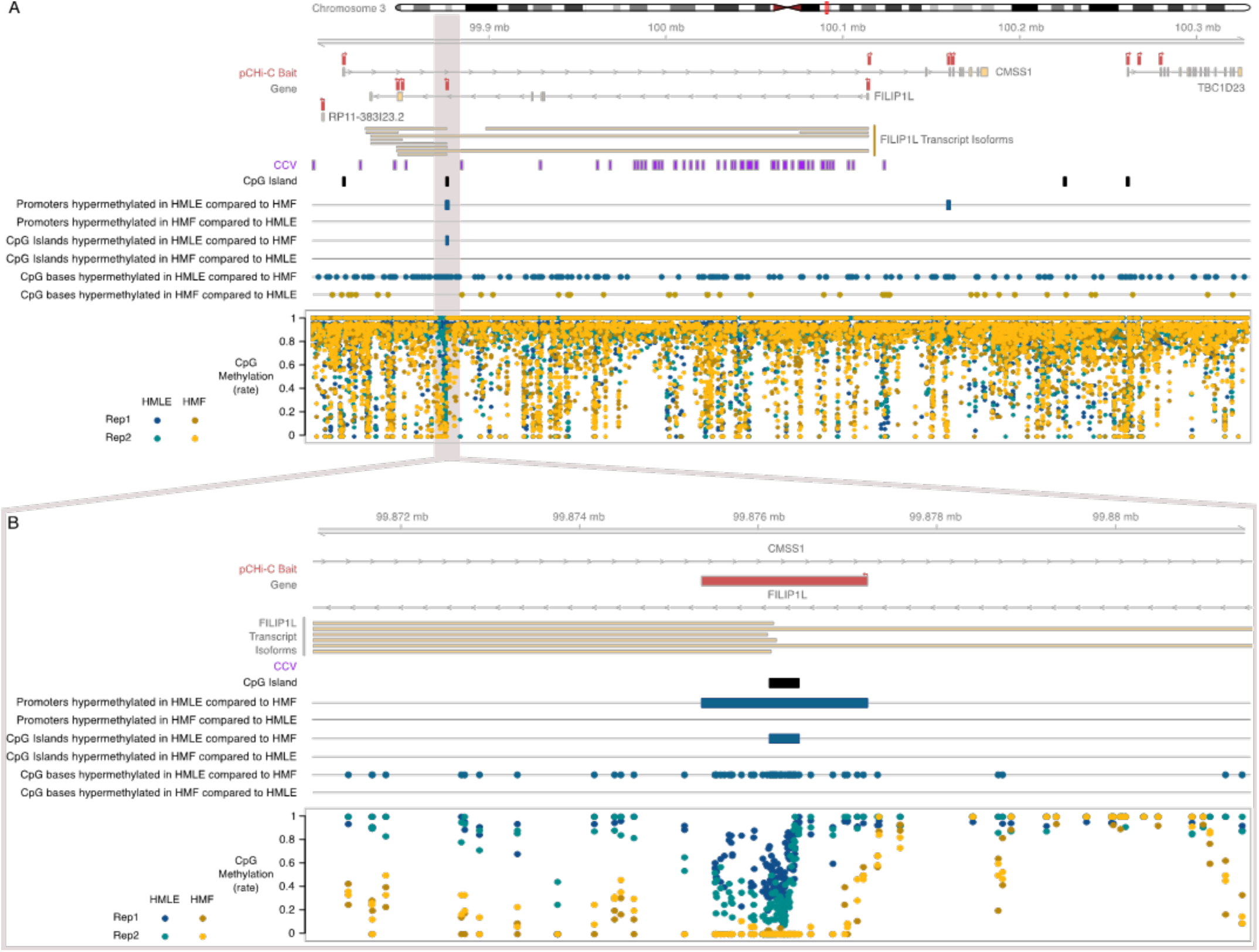
Differentially methylated CpG bases, CpG islands and promoters at the 3q12.1-FILIP1L locus. (A) Methylation rate (0 to 1) for individual CpG bases are shown for two HMLE (blue and teal) and two HMF (yellow and brown) biological replicates aligned with Ensembl genes (grey/yellow), pCHi-C baits (red), CCVs (purple) and CpG islands (black) at the 3q12.1-*FILIP1L* locus. Differentially methylated CpG bases, CpG islands and gene promoter bins are shown directly above the methylation rates. (B) An ∼10kb region at chr3:99,871,000-99,881,000 encompassing the *FILIP1L* P3 promoter bin (chr3:99875365-99877224) is shown at higher resolution. This promoter bin, which aligns with a CpG island (chr3:99876126-99876457) is hypermethylated in HMLE compared to HMF.

We were unable to differentiate between P3- and P2-transcribed *FILIP1L* isoforms using qPCR. Accordingly, we used Western blotting to further examine isoform-specific expression of *FILIP1L.* We first transfected T-47D, in which *FILIP1L* expression is undetectable (https://www.proteinatlas.org), with cDNA clones of two *FILIP1L* isoforms – ENST00000487087 and ENST00000383694 transcribed from P2 and P3, respectively, and observed bands at ∼90 kDa (ENST00000487087, FILIP1L_P2) and ∼120 kDa (ENST00000383694, FILIP1L_P3; Figure 5A). We were unable to detect the P2-transcribed isoform (∼90 kDa band) in any of the primary cells or cell lines (Figure 5B). We detected P3-transcribed isoforms (∼120 kDa) in HMLE but not in MCF10A or non-transfected T-47D precluding further investigation of expression of these isoforms in an epithelial cell line model; they were, however, detected in both HMF and GS2.

**Figure 5.**
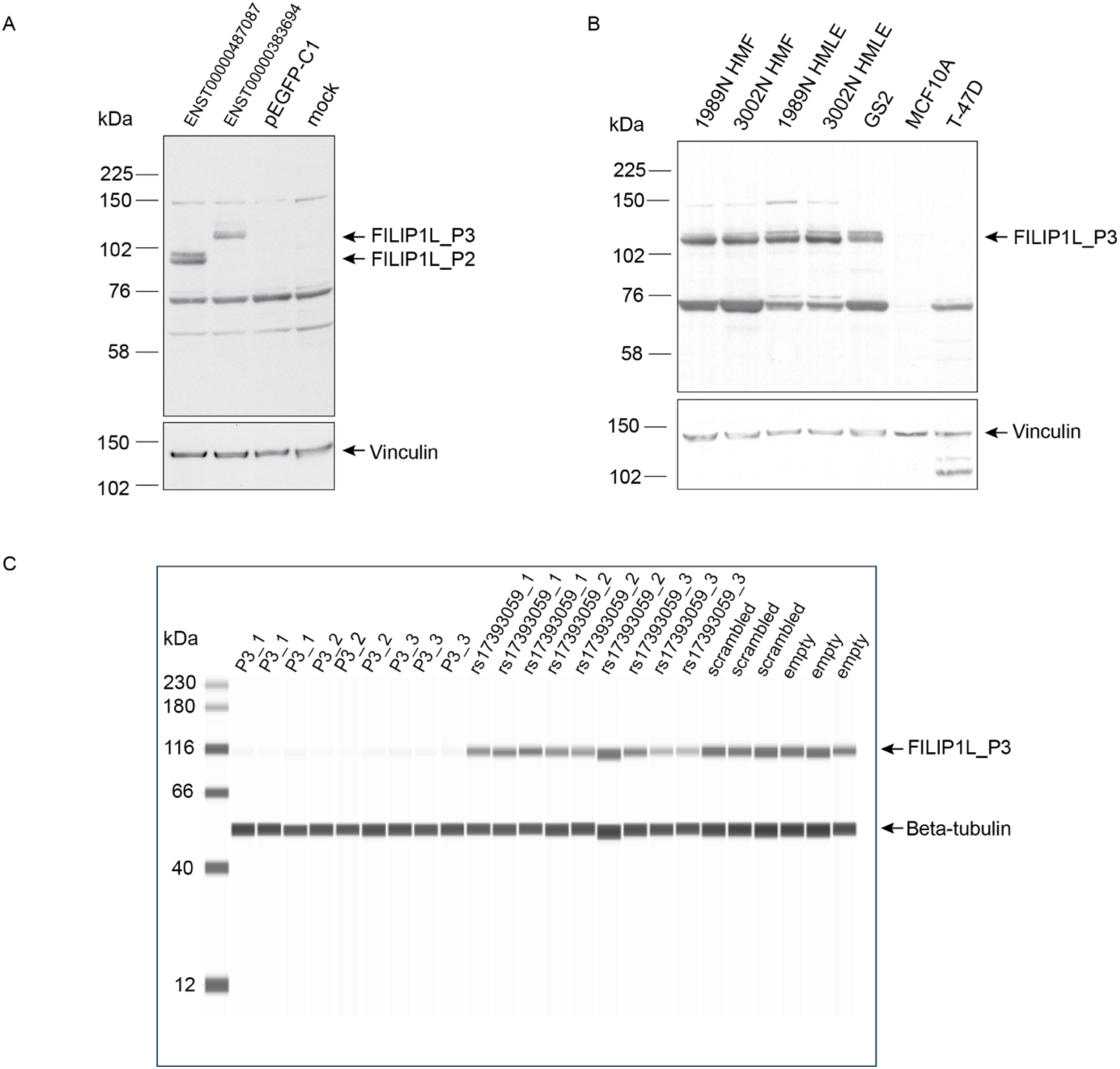
Western blots of FILIP1L in primary cells and cell lines and CRISPRi experiments. (A) Western blots of whole cell lysates from T-47D (which does not express endogenous *FILIP1L*) transfected with cDNA of a *FILIP1L* isoform that is transcribed from the P3 promoter bin (ENST00000383694) and the P2 promoter bin (ENST00000487087) demonstrating protein bands of ∼120 kDa and 90 kDa, respectively. Blots were probed with an anti-FILIP1L antibody and also show a non-specific band at ∼70 kDa which is present in all samples. (B) HMF, HMLE and GS2 whole cell lysates show protein bands corresponding to the *FILIP1L* isoforms transcribed from P3 but not P2. Consistent with RNA-seq data generated by The Human Protein Atlas project (https://www.proteinatlas.org) there are no FILIP1L-specific bands in the MCF10A or T-47D extracts. (C) CRISPRi in GS2 with three sgRNAs targeting the P3 promoter bin (P3_1, P3_2, P3_3), three sgRNAs targeting rs17393059 (rs17393059_1, rs17393059_2, rs17393059_3), one non-targeting scrambled sgRNA (scrambled), and no sgRNA (empty vector) (each performed in triplicate). Whole cell lysates were run on the JESS Simple Western™ instrument. As a loading control, blots were probed with an antibody against vinculin (Figure 5A, 5B) or beta tubulin (Figure 5C)

To determine directly whether rs17393059 might impact *FILIP1L* expression in fibroblasts, we used CRISPR interference (CRISPRi)^32^. We designed single guide RNAs (sgRNAs) targeting rs17393059, the P3 promoter bin (positive control), the index SNP at this locus (rs506186), the P2 promoter bin and a non-targeting scrambled sgRNA (negative controls; Supplementary Table 6). We then introduced these guides into GS2 cells that expressed dCas9-KRAB and carried out RNA-seq analysis; we found that targeting the P3 promoter and rs17393059 reduced expression of *FILIP1L* by 40% (P3 promoter) and 23 to 31% (rs17393059; Table 1) but not the expression of neighbouring genes (Supplementary Table 7). To determine whether this reduction in expression of *FILIP1L* results in a reduction in the abundance of FILIP1L protein we repeated the CRISPRi experiment with sgRNAs targeting the P3 promoter bin (three guides), rs17393059 (three guides) and the non-targeting scrambled sgRNA and measured FILIP1L protein levels using the Jess^TM^ Simple Western system. All three guides targeting the P3 promoter resulted in a reduction in FILIP1L of approximately 80%; two of the guides targeting rs17393059 reduced expression of FILIP1L, and for one of these, the reduction was significant (guide 2: -16%, *P* = 0.11, guide 3: -26% *P* = 0.01; Figure 5C, Supplementary Table 8). In summary, these data provide strong evidence of an association between genotype and *FILIP1L* expression in fibroblasts and implicate rs17393059 as a potentially functional variant that influences expression from a *FILIP1L* promoter that is preferentially active in HMF.

**Table 1:** Expression of *FILIP1L* versus no sgRNA in GS2 CRISPRi.

|  | *FILIP1L* |  |  |
| --- | --- | --- | --- |
|  | Log<sub>2</sub> FC | *P* | *P*<sub>FDR</sub><sup>1</sup> |
| rs17393059_guide1 | -0.37 | 0.03 | 0.46 |
| rs17393059_guide2 | -0.53 | 0.004 | 0.09 |
| rs17393059_guide3 | -0.46 | 0.01 | 0.17 |
| P3 promoter_guide1 | -0.74 | 0.0002 | 0.02 |
| P2 promoter | -0.23 | 0.18 | 0.85 |
| rs506186 | -0.21 | 0.21 | 0.74 |
| scrambled | 0.05 | 0.77 | 1.00 |

<sup>1</sup>an FDR corrected *P*-value <0.1 was considered significant

## Discussion

GWAS have transformed our understanding of the genetic architecture of breast cancer risk, however translating these findings into a greater understanding of the mechanisms that influence the risk of breast cancer will require the identification of functional variants and the targets of these functional variants. By capturing the physical interactions that occur between regulatory elements and the genes that they regulate, CHi-C has the potential to link variants to genes. Aligning the large number of CCVs that are the product of fine scale mapping with epigenetic markers that correlate with regulatory activity can prioritise potentially functional CCVs and infer the cell type in which they influence risk (reviewed in ^13^)

Previous CHi-C analyses of breast cancer GWAS risk loci have been carried out^21,24,33^. These studies have focused on breast cancer or “normal” immortalised breast epithelial cell lines for library generation and used a restriction-enzyme (HindIII) for chromatin fragmentation. Despite the evidence demonstrating the role of the stroma in cancer predisposition, progression and metastasis^4,34^, to our knowledge this is the first study to include data generated in fibroblasts for the functional follow up of breast cancer GWAS loci. By using the Dovetail Omni-C Kit for library preparation we have been able to reduce the input number of cells by more than five-fold and generate libraries in primary cells as well as cell lines. While cell lines remain essential for carrying out functional assays such as CRISPRi, by including primary cells in our analyses and only selecting interactions that are observed in a cell line and replicated in a primary cell, we have tried to ensure that our results are relevant to the “at risk” breast in which breast cancers arise. By generating genome-wide CHi-C data, capturing annotated gene promoters rather than GWAS linkage disequilibrium blocks, along with genome-wide epigenetic data in the same primary cells and cell lines, we have expanded the resources that are available to breast cancer researchers and addressed a relative lack of such data generated in mammary fibroblasts.

We used a strategy of focusing on interaction peaks that were identified in a fibroblast cell line and replicated in primary fibroblasts, or identified in an epithelial cell line and replicated in primary epithelial cells, but not in both. Using this strategy we have incidentally identified plausible target genes and potentially functional variants at 8q23.3 (*TRPS1*) and 2q14.2 (*INHBB*) which would be predicted to act in a cell autonomous manner to influence breast cancer risk. Further studies will be required to determine whether our criteria have correctly prioritized these variants over the other CCVs at these loci.

The primary aim of this work was, however, to determine whether perturbation of expression of a GWAS target gene in mammary fibroblasts, rather than epithelial cells, might impact breast cancer risk. We found evidence that this might be the case at the 3q12.1-*FILIP1L* locus which, to our knowledge has not been investigated previously. GTEx eQTL data provide compelling evidence for an association between at least one 3q12.1 CCV and expression of *FILIP1L* in cultured fibroblasts. Genome-wide functional data demonstrate preferential expression of P3-transcribed *FILIP1L* isoforms in fibroblasts and support rs17393059 as a potentially functional variant; directly targeting rs17393059 using CRISPRi confirmed a modest functional association between rs17393059, *FILIP1L* mRNA expression and FILIP1L protein levels in immortalised mammary fibroblasts.

*FILIP1L*, formerly named deleted in ovarian cancer (*DOC1*), has previously been reported to act as a tumour suppressor gene in epithelial cancers including ovarian, breast, lung, colon, pancreatic and prostate cancer^27–29^. *FILIP1L* expression has been shown to inhibit WNT/β-catenin signalling, epithelial-to-mesenchymal transition, metastasis and chemoresistance in ovarian cancer, and was downregulated in a series of cancer cells compared with their normal equivalent epithelial cells^27,35^. The SNPs that annotate GWAS signals are enriched in DNA sequences that carry the hallmarks of active regulatory regions and this enrichment has been shown to be cell-selective within physiologically relevant cell types suggesting a tissue-specific regulatory role for many common variants ^12,36,37^. However, there was no evidence of an association between 3q12.1 variants and *FILIP1L* expression in (predominantly epithelial) mammary tissue in GTEx data and it has been reported that the down-regulation of *FILIP1L* in epithelial cancer cells occurs predominantly via promoter methylation^27–29^. These data raise the possibility that breast cancer risk might be impacted by regulation of expression of *FILIP1L* in more than one cell type by more than one mechanism, with epigenetic regulation (promoter methylation) influencing expression in epithelial cells and genetic regulation (perturbation of enhancer activity by a common variant) influencing expression in fibroblasts. Consistent with *FILIP1L* also acting as a tumour suppressor in fibroblasts, in the GTEx eQTL analysis the major (A) allele of rs17393059 was associated with higher levels of *FILIP1L* expression in fibroblasts and lower levels of estrogen receptor-positive breast cancer risk ^10^. However, further work will be needed to unravel the mechanism(s) by which *FILP1L* expression in fibroblasts might impact the behaviour of epithelial cells *in trans* to reduce risk. In conclusion, we have demonstrated that rs17393059, which maps to the 3q12.1 breast cancer risk locus, perturbs expression of *FILIP1L* in mammary fibroblasts. Unravelling the exact mechanism(s) by which this locus influences breast cancer risk, however, will require additional work.

## Data Availability

Raw capture pCHi-C (HMLE x2, HMF x2, MCF10A x3, T-47D x2, GS2 x2), H3K27ac CUT&Tag (MCF10A x2, T-47D x2, GS2 x2, HMLE x2, HMF x2),
whole genome NEBNext EmSeq (HMLE x2, HMF x2), bulk RNA-seq (HMLE x5, HMF x4,) and CRISPRi-modified RNA-seq (GS2 x21) generated in this study are deposited in the Sequence Read Archive under the submission number PRJNA1235642. Processed pCHi-C interaction peaks (HMLE x1, HMF x1, MCF10A x1, T-47D x1, GS2 x1) generated in this study are deposited in the Gene Expression Omnibus under the submission number GSE291943. Processed pCHi-C interaction peaks can be uploaded to the WashU Epigenome Browser (https://epigenomegateway.wustl.edu/) in the Tracks function by selecting Local text tracks and then the Long range format by CHiCANE option from the Choose text file type drop
down menu. CHiCANE interaction peak files must be unzipped prior to WashU upload; the WashU browser will accept zipped files, but they will appear to contain no interactions. Processed H3K27ac CUT&Tag consensus peaks (HMLE x1, HMF x1, MCF10A x1, T-47D x1,GS2 x1) generated in this study are deposited in the Gene Expression Omnibus under the submission number GSE291944. The Genotype-Tissue Expression (GTEx) Project was supported by the Common Fund (https://commonfund.nih.gov/GTEx) of the Office of the
Director of the National Institutes of Health and by NCI, NHGRI, NHLBI, NIDA, NIMH, and NINDS. The data used for the analyses described in this manuscript were obtained from the GTEx Portal on 01/25/2024.

https://www.encodeproject.org

---

**Supplementary Figure 1:**
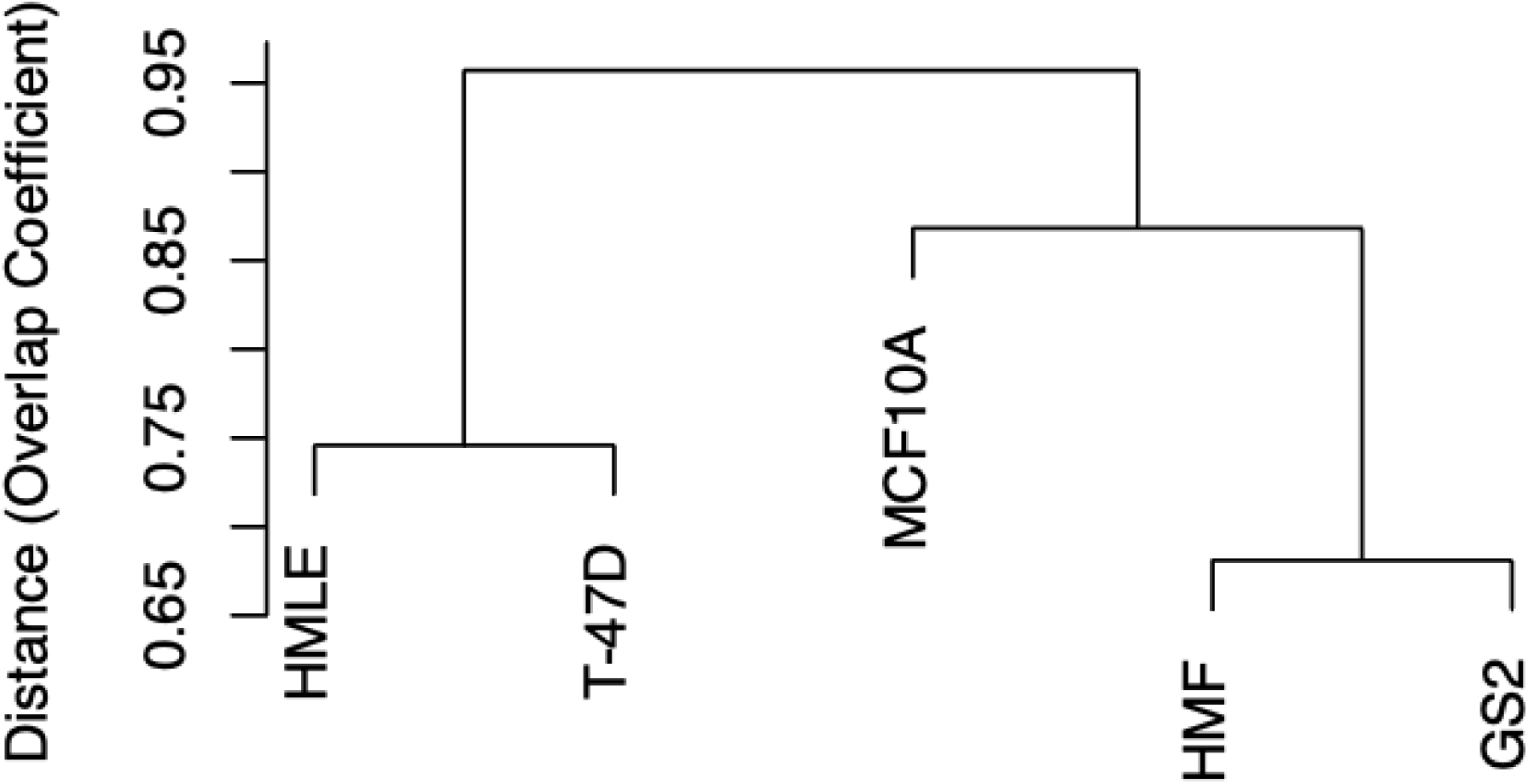
Dendrogram of the total number and cell type-specificity of interaction peaks in pCHi-C libraries generated in epithelial cells (HMLE, MCF10A and T-47D) and fibroblasts (HMF and GS2). **Legend:** Dendrogram showing the distance between epithelial cells (HMLE, MCF10A and T-47D) and fibroblasts (HMF and GS2), calculated by overlap coefficient. Distances are between 1.0 and 0 with smaller distances indicating greater similarity.

**Supplementary Figure 2:**
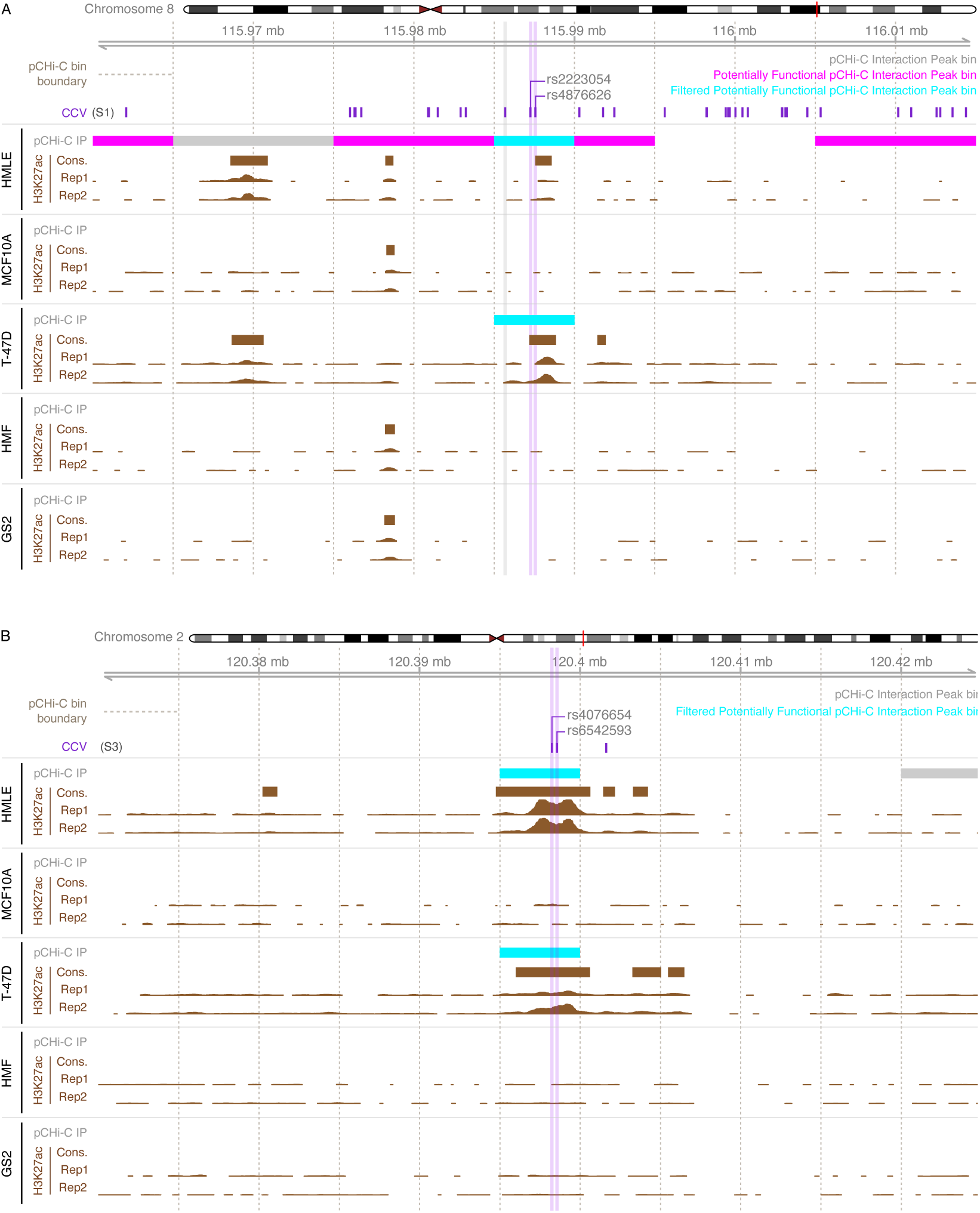
Zoomed in view of interaction peaks at (A) 8q23.3-*TRPS1* and (B) 2q14.2-*INHBB* in epithelial cell and fibroblast pCHi-C libraries. **Legend**: Uncaptured ends of potentially functional interaction peaks are in pink and the subset that are unique to epithelial cells, and at which the CCVs also colocalize with an H3K27ac peak in at least one epithelial cell type, are in turquoise.

**Supplementary Figure 3:**
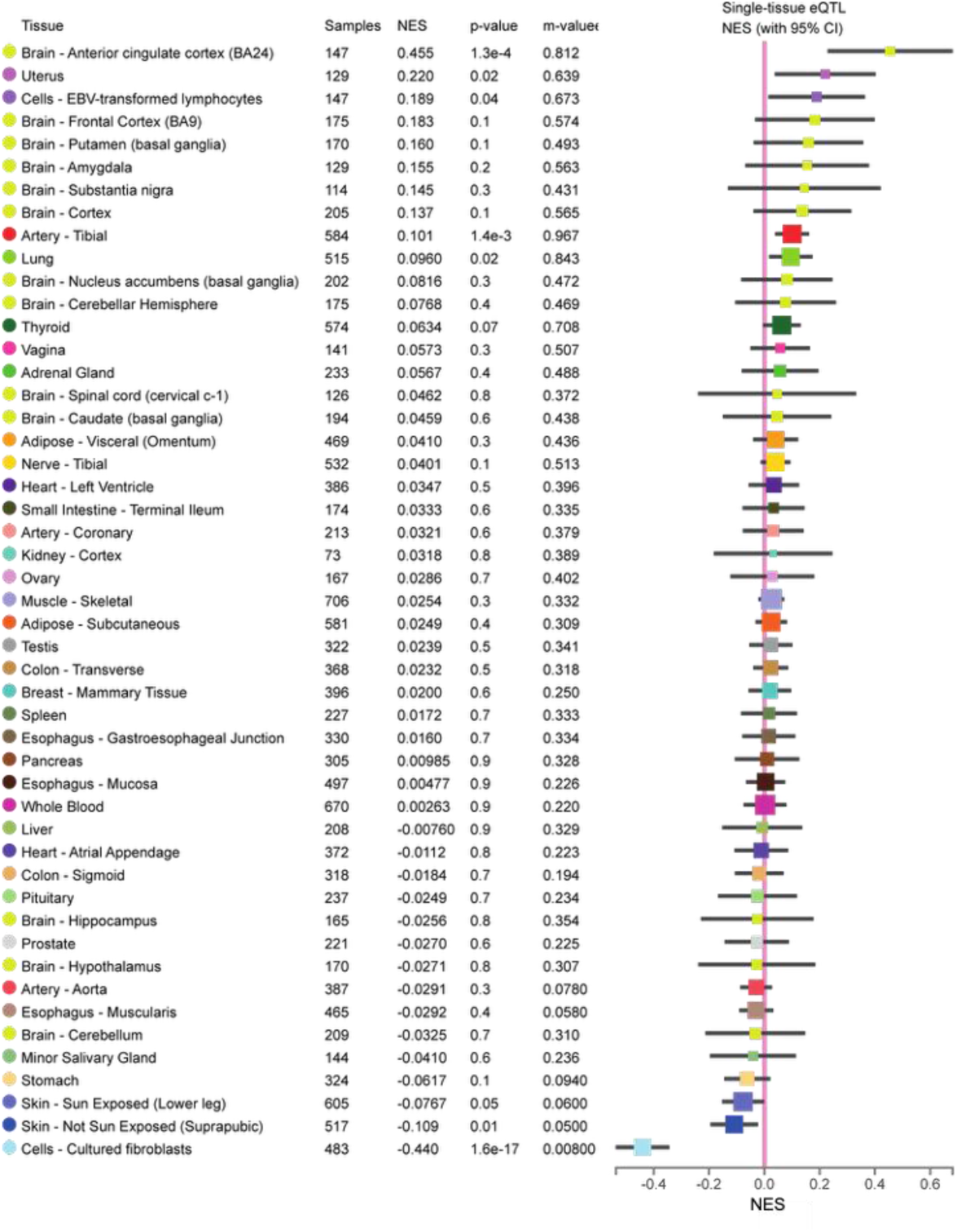
Multi-tissue eQTL comparison of rs17393059 and *FILIP1L* expression. **Legend:** Normalised expression values (NES) based on quantile normalisation within each tissue followed by inverse quantile normalisation for each gene across samples are shown for *FILIP1L* and rs17393059. The figure was downloaded from the GTEx data portal at https://gtexportal.org. The number of samples that were included in the analysis (Samples), the *p*-value from a *t*-test that compares the observed NES from single-tissue eQTL analysis to a null NES of 0 (*p*-value) and the posterior probability that an eQTL effect exists in each tissue tested in the cross-tissue meta-analysis (m-value[44, 45]) are indicated. The size of the box is proportional to the inverse variance of the NES estimate and horizontal lines represent 95% confidence intervals.

**Supplementary Figure 4:**
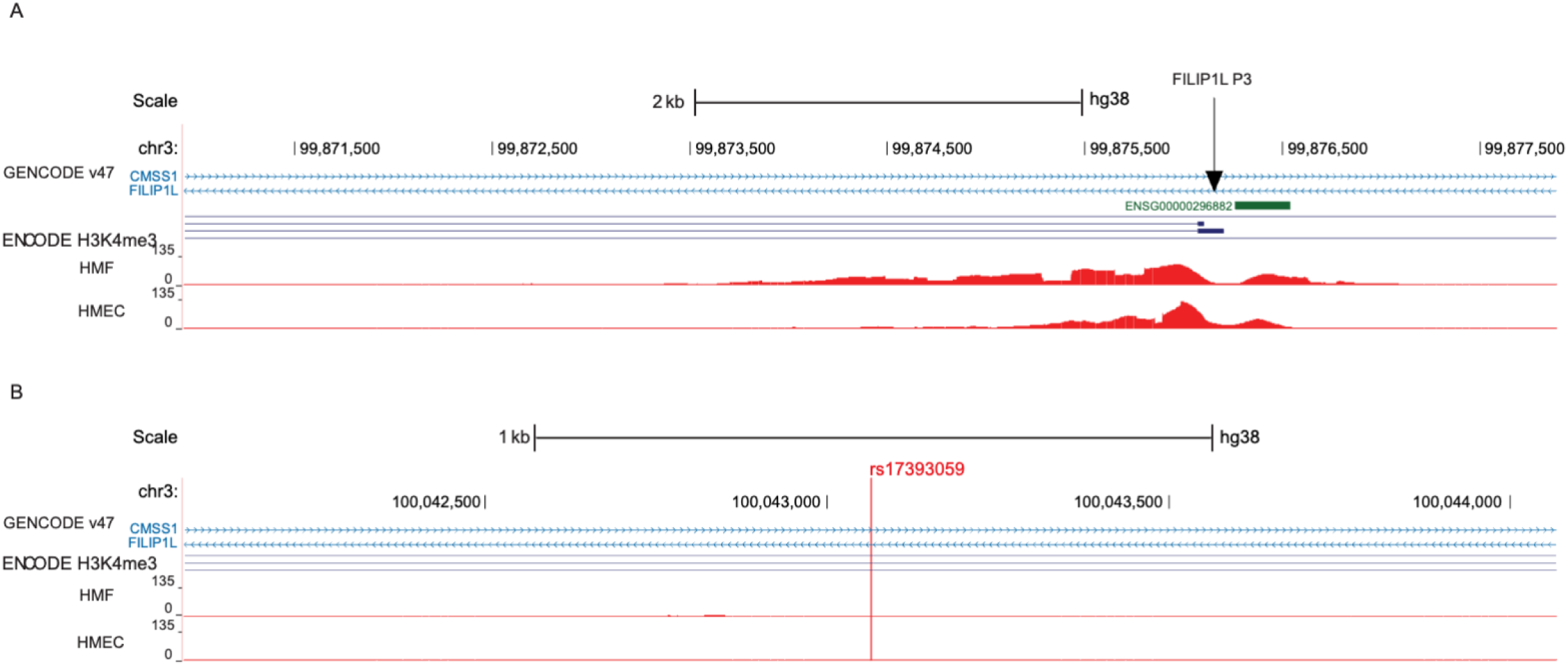
Extended H3K4me3 histone modification is present at the *FILIP1L* P3 promoter locus (A) but is absent at rs17393059 (B). **Legend:** H3K4me3 signals generated in HMF and HMEC as part of the ENCODE project (https://www.encodeproject.org/).

**Supplementary Figure 5:**
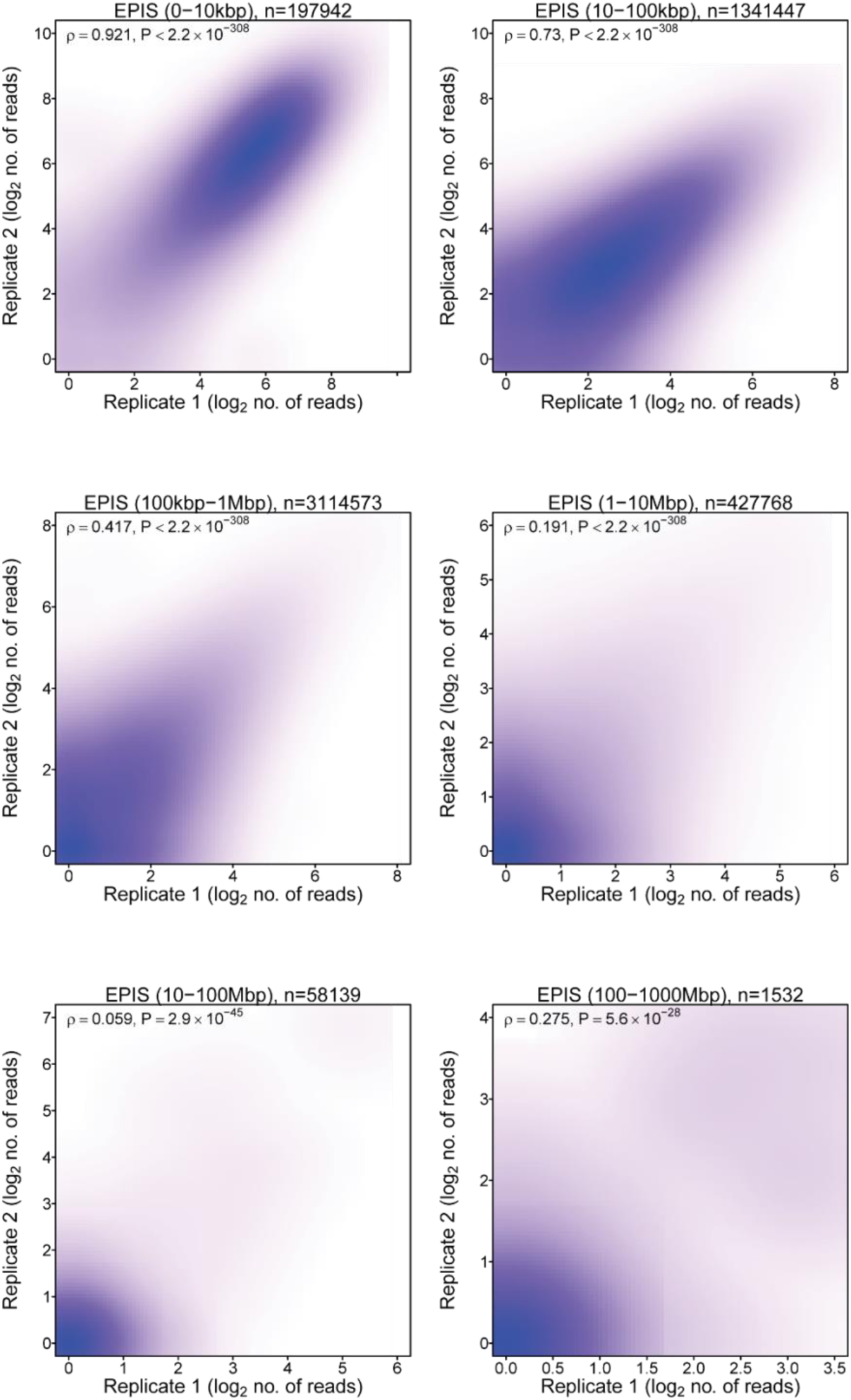
Scatterplots showing the correlation between duplicate HMLE pCHi-C libraries. **Legend**: Scatterplot showing the association (quantified as Spearman’s *ρ*) between duplicate HMLE libraries based on the number of raw di-tags mapping to each combination of captured promoter bin and non-captured “target” bins.

**Supplementary Figure 6:**
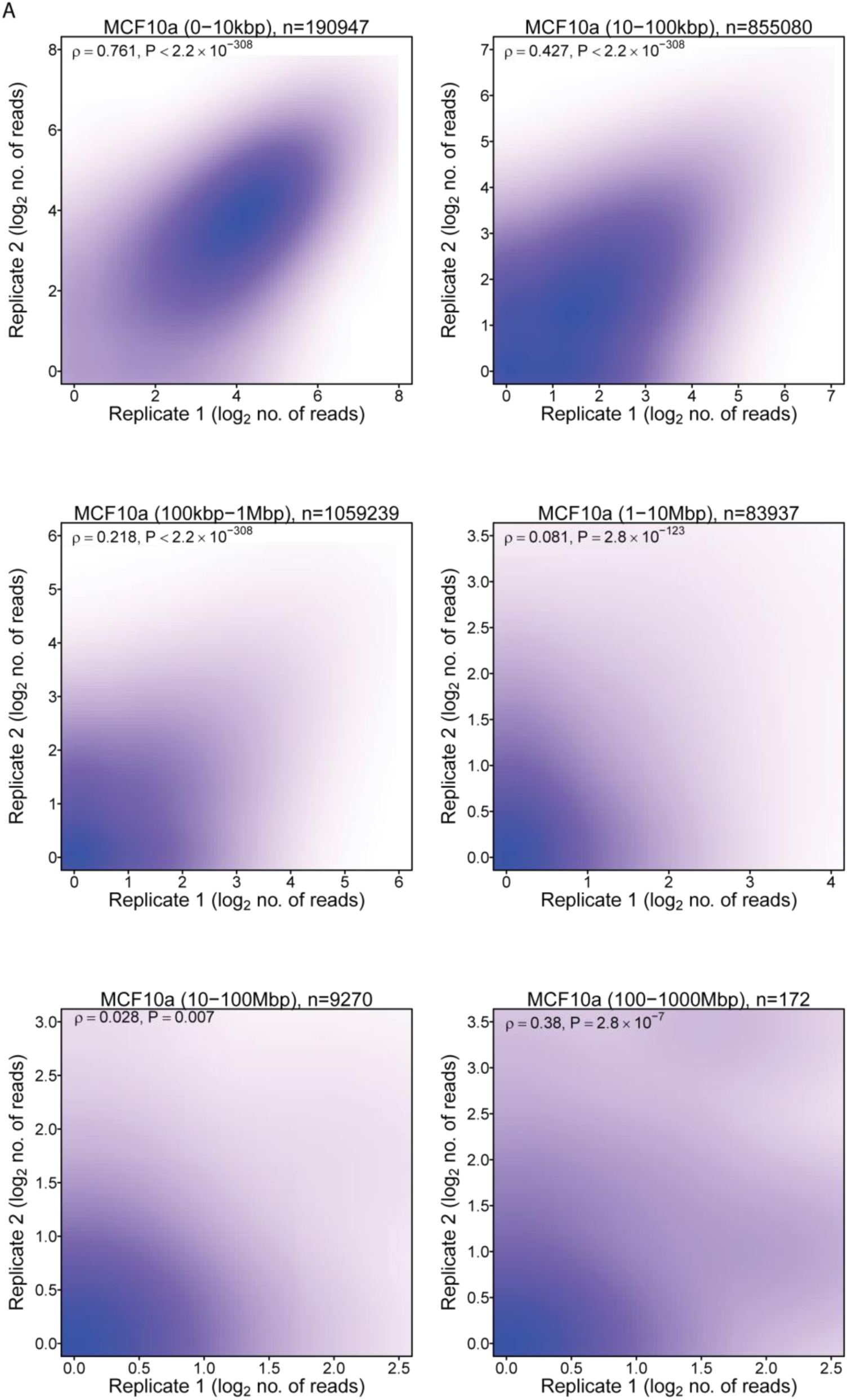
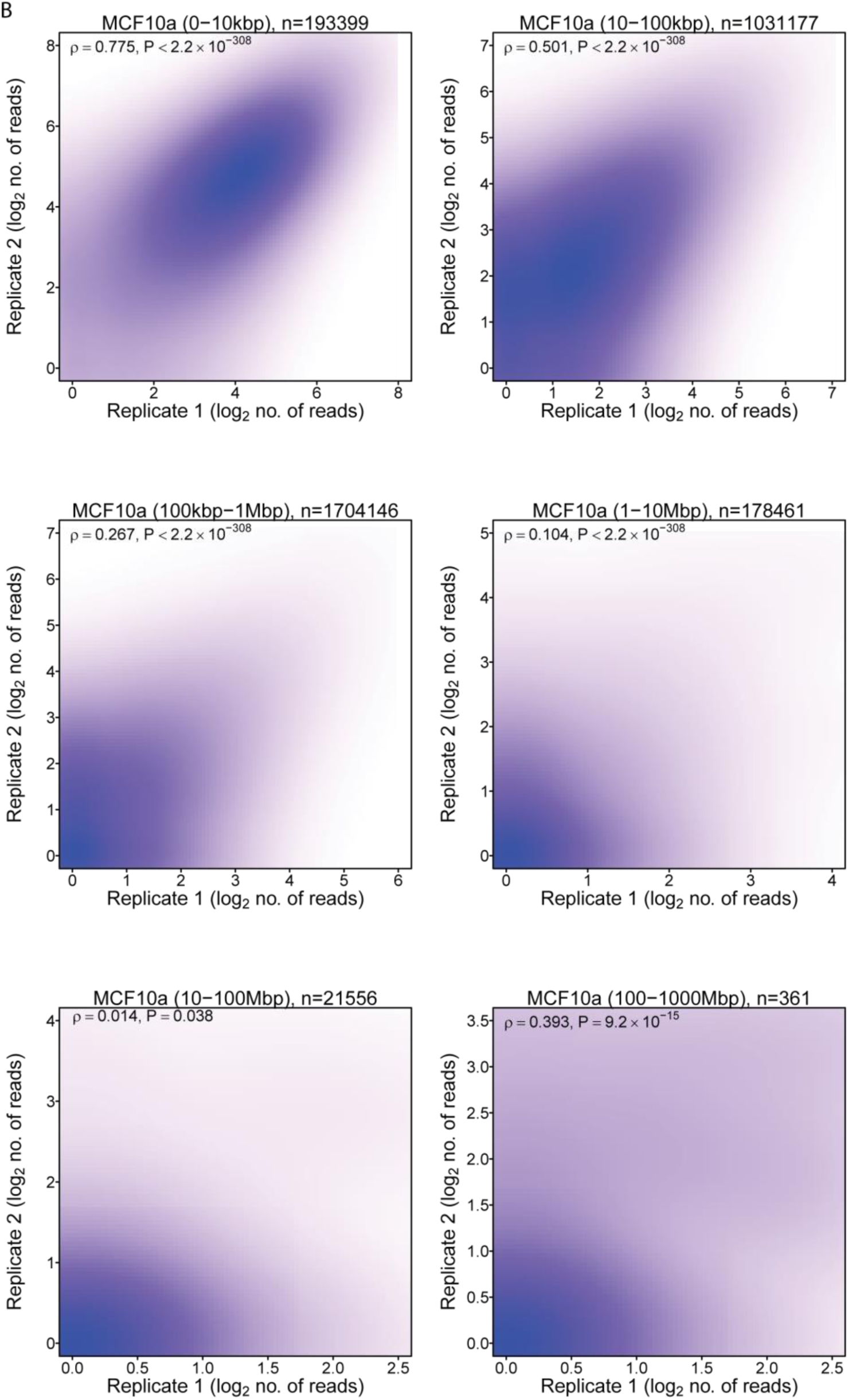
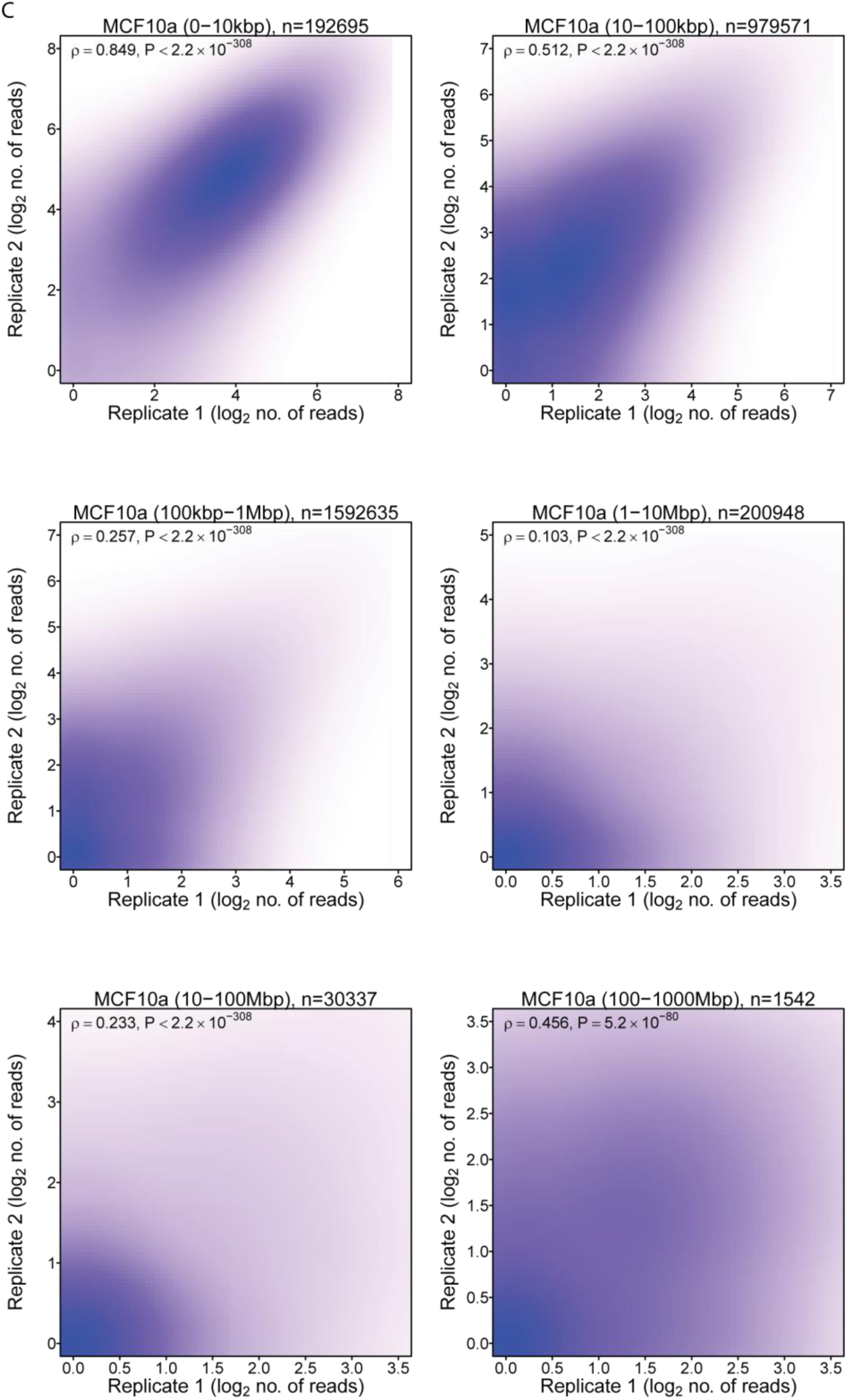
Scatterplots showing the correlation between triplicate MCF10A pCHi-C libraries. **Legend**: Scatterplot showing the association (quantified as Spearman’s *ρ*) between triplicate MCF10A libraries (A) A vs B, (B) A vs C, (C) B vs C based on the number of raw di-tags mapping to each combination of captured promoter bin and non-captured “target” bins.

**Supplementary Figure 7:**
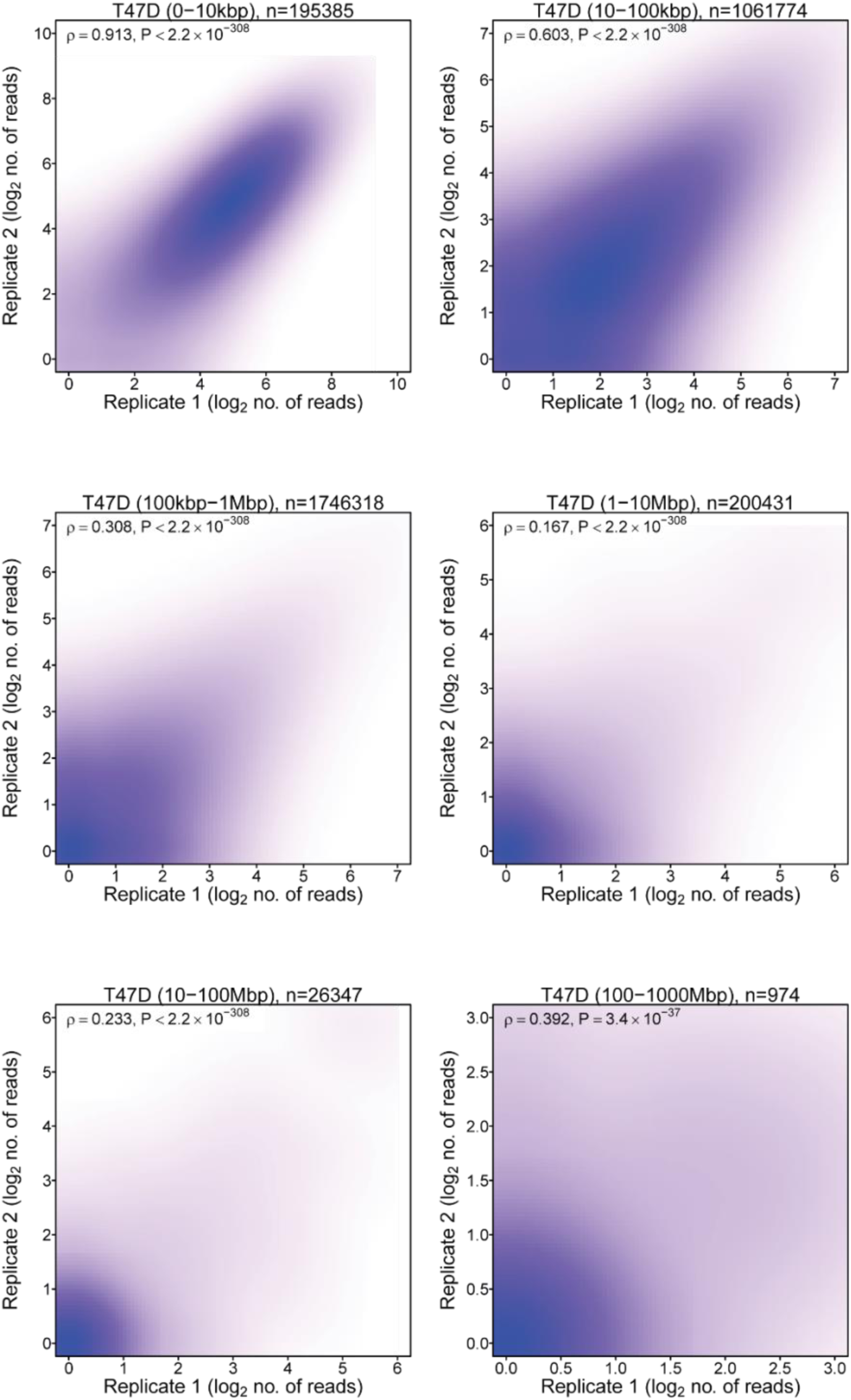
Scatterplots showing the correlation between duplicate T-47D pCHi-C libraries. **Legend**: Scatterplot showing the association (quantified as Spearman’s *ρ*) between duplicate T-47D libraries based on the number of raw di-tags mapping to each combination of captured promoter bin and non-captured “target” bins.

**Supplementary Figure 8:**
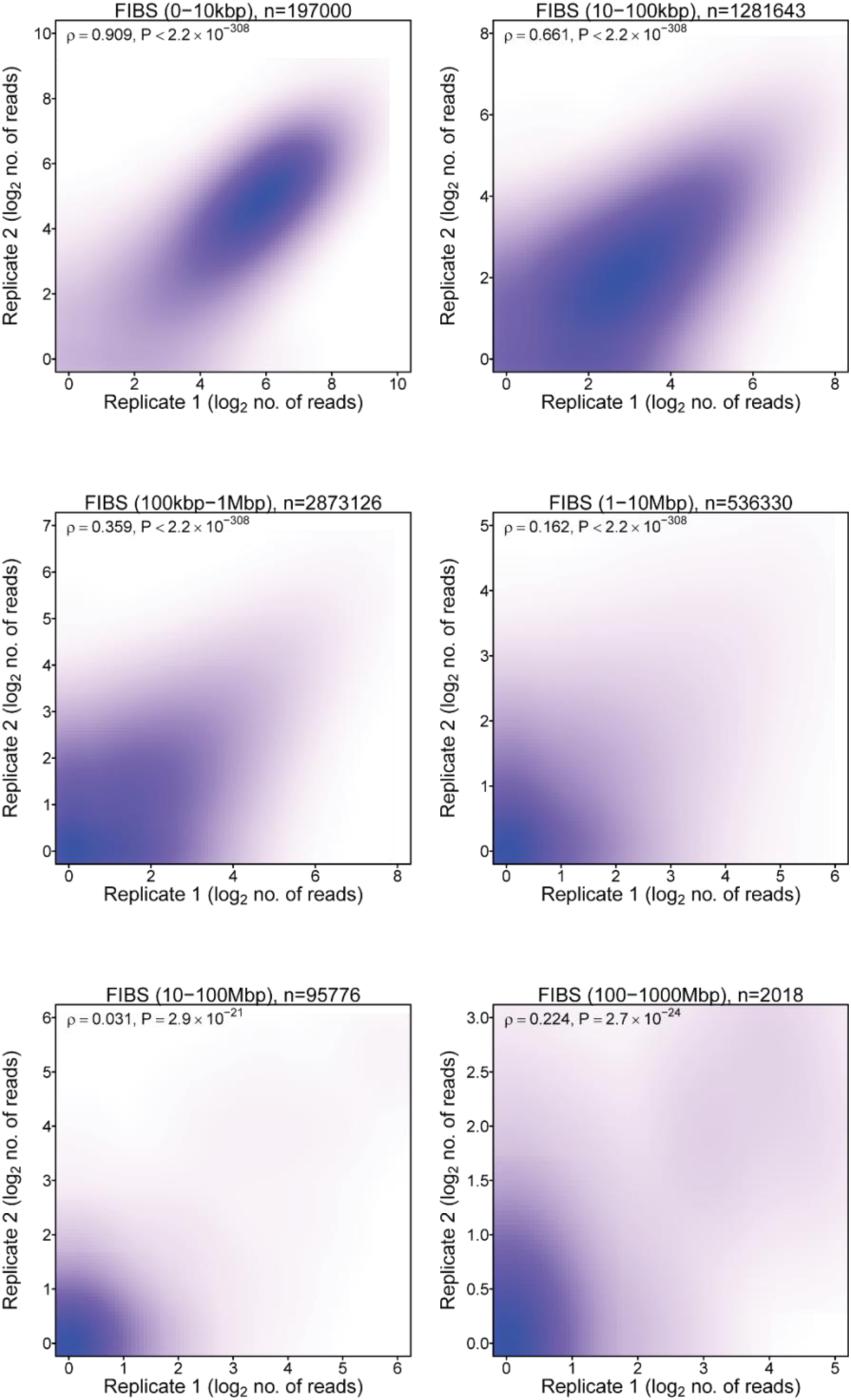
Scatterplots showing the correlation between duplicate HMF pCHi-C libraries. **Legend**: Scatterplot showing the association (quantified as Spearman’s *ρ*) between duplicate HMF libraries based on the number of raw di-tags mapping to each combination of captured promoter bin and non-captured “target” bins.

**Supplementary Figure 9:**
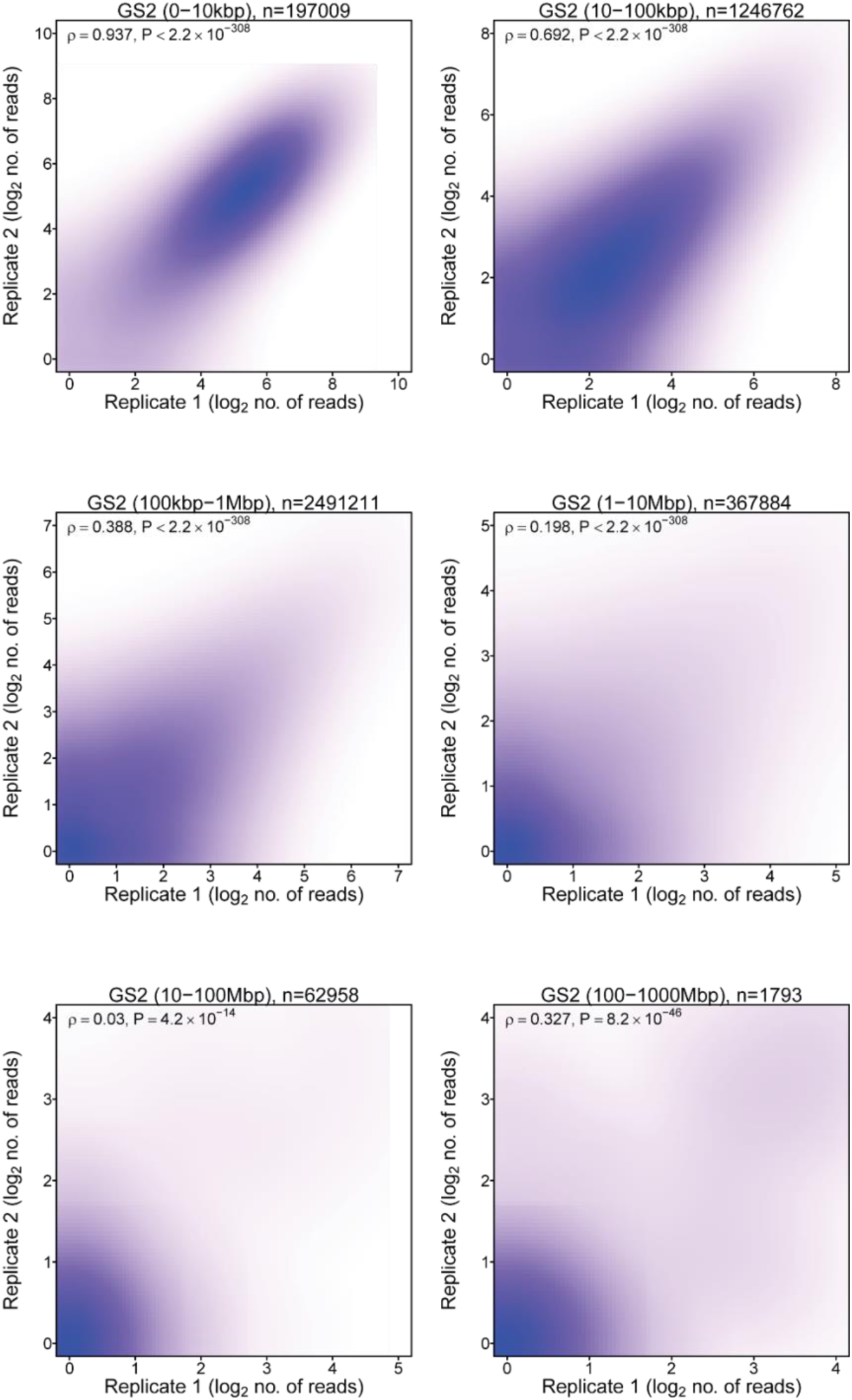
Scatterplots showing the correlation between duplicate GS2 pCHi-C libraries. **Legend**: Scatterplot showing the association (quantified as Spearman’s *ρ*) between duplicate GS2 libraries based on the number of raw di-tags mapping to each combination of captured promoter bin and non-captured “target” bins.

**Supplementary Figure 10:**
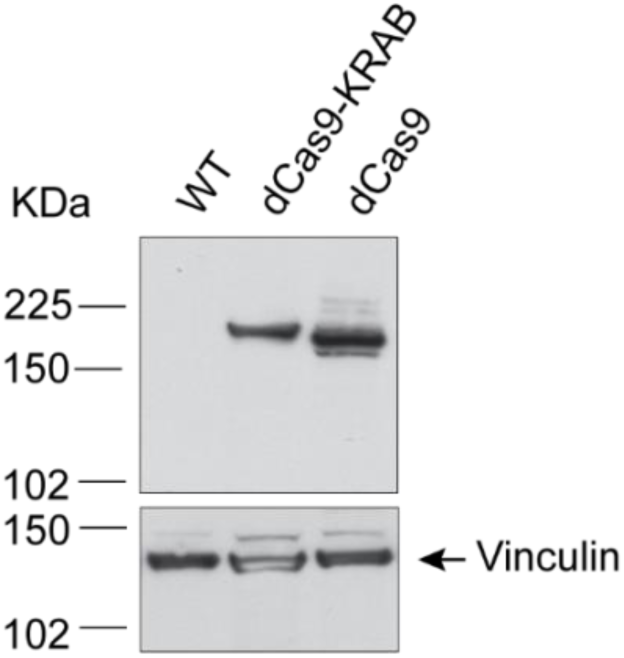
Western blot using anti-dCas9 antibody in parental GS2 and GS2 that have been modified to express dCas9-KRAB and dCas9. **Legend**: Western blot of whole cell lysates from parental GS2 (Lane 1: WT, negative control) and GS2 that have been modified to express dCas9-KRAB (Lane 2) and dCas9 (Lane 3: positive control). Blots were probed with an anti-dCas9 antibody. As a loading control, blots were probed with an antibody against vinculin.

## Methods

### Cell line culture

T-47D cells (HTB-133, ATCC) were cultured in RPMI-1640 medium (Gibco) supplemented with 10% foetal bovine serum (FBS; Gibco), 10μg/ml human insulin (Sigma) and 1X penicillin-streptomycin (Sigma) at 37**°**C, 5% CO_2_. MCF10A (CRL-10317, ATCC) were cultured in DMEM:F12 medium (Invitrogen) supplemented with 5% horse serum (Invitrogen), 20ng/ml epidermal growth factor (PeproTech), 0.5mg/ml hydrocortisone (Sigma), 100ng/ml Cholera toxin (Sigma), 10µg/ml bovine insulin (Sigma), 1X penicillin-streptomycin (Invitrogen) at 37**°**C, 5% CO_2_. GS2 cells, which are hTERT immortalised human mammary fibroblasts (kindly provided by Professor Clare Isacke; The Institute of Cancer Research, London, UK) were cultured in DMEM media (Gibco) supplemented with 10% FBS at 37**°**C, 10% CO_2_. 293T cells (CRL-3216, ATCC) were cultured in DMEM media (Gibco) supplemented with 10% FBS at 37**°**C, 5% CO_2_. All cell lines were regularly tested for *Mycoplasma* contamination.

### Primary cells derivation and culture

#### Tissue preparation

On arrival at the Breast Cancer Now Tissue Bank, tissue was washed once with ethanol and then three times with RPMI-1640 media (Sigma) containing 25mM HEPES supplemented with 5% FBS (Gibco), 100U/ml penicillin, 0.1mg/ml streptomycin (pen/strep; Sigma), 5µg/ml amphotericin-B (Sigma) and 20μg/ml Gentamicin (Sigma).

Tissue was chopped into small pieces and digested for 12 to 16 hours at 37°C on a rotary shaker in RPMI-1640 medium containing 25mM HEPES, supplemented with 5% FBS, pen/strep and 5µg/ml amphotericin-B containing 1mg/ml collagenase 1A (Sigma) and hyaluronidase (Sigma). The digested tissue was centrifuged at 380g for 20 minutes and washed in medium three times to remove enzymes. The tissue isolates were then sedimented three times at 1g for 30 minutes to collect the denser ductal tree fragments (DTF) (ductal tree containing terminal duct lobular units and ducts). The DTF are centrifuged (380g, 3minutes) and frozen for specific cell isolation. The supernatants containing the fibroblasts were centrifuged (380g, 3 minutes) and the cell pellets were re-suspended and cultured in DMEM:F12 (Sigma) supplemented with 10% FBS, 0.5μg/ml hydrocortisone (Sigma), 10μg/ml apo-transferrin (Sigma), 5μg/ml insulin (Sigma), 10ng/ml EGF (Sigma), pen/strep and 2.5μg/ml amphotericin-B (Breast Culture Medium, BCM).

#### Isolation and expansion of human mammary luminal epithelial cells (HMLE)

HMLE were obtained from two women (anonymised identifiers 1989, 3002) undergoing reduction mammoplasty. DTF were digested using 0.25% trypsin/0.1% EDTA (HyClone SV30031.01, Fisher Scientific) and DNase I (Merck) was added to ensure the cells were single. Cells were counted before adding fluorescently conjugated antibodies to EPCAM and CD10 and then sorted using Fluorescence Activated Cell Sorting (FACS) to separate the EPCAM+ cells (luminal epithelial cells), CD10+ cells (myoepithelial cells) and double negatives (microenvironment cells). The fraction containing epithelial cells was then centrifuged (380g, 3minutes) and the cell pellet re-suspended and cultured in BCM, on collagen-coated tissue culture plates. HMLE were grown for 1 passage before being frozen down and stored in liquid nitrogen. For this work, vials of 300,000 passage 1 HMLE were thawed and seeded into 6-well tissue culture plates. HMLE were expanded in BCM to achieve final passage 2 numbers of 5x10^6^ (pCHi-C libraries), 1x10^6^ (RNA-seq), 2x10^5^ (H3K27ac CUT&Tag), 5x10^6^ (Western blotting) and 1x10^6^ (NEBNext EmSeq).

#### Isolation and expansion of human mammary fibroblasts (HMF)

HMF were obtained from two women (anonymised identifiers 1989, 3002) undergoing reduction mammoplasty. Explants were chopped to small pieces and left to attach to flasks in BCM. Cells then grew from the explants, these were mostly fibroblasts, but limited numbers of epithelial cells were present. Cells of epithelial origin do not survive well with passaging or freezing and so taking the cells through 2 passages in BCM and then a further passage in DMEM:F12 (Sigma) supplemented with 10% FBS, 100U/ml penicillin, 0.1mg/ml streptomycin, 2.5μg/ml amphotericin-B (Fibroblast Media, FM) meant epithelial cells could no longer be identified. Remaining cells were frozen at 1 million cells per vial. For this work, vials of 1x10^6^ fibroblasts were thawed and seeded into T75 tissue culture flasks. Fibroblasts were expanded in DMEM media (Gibco) supplemented with 10% FBS to achieve final numbers of passage 3 HMF for pCHi-C, RNA-seq, H3K27ac CUT&Tag, Western blotting and NEBNext EmSeq as above.

#### Dovetail Omni-C library generation

Omni-C libraries were generated using the Dovetail Omni-C Kit according to the manufacturer’s instructions (Dovetail Genomics, 21005). Briefly, 1 million cells (HMLE, MCF10A, T-47D, HMF and GS2) were fixed with a combination of formaldehyde and disuccinimidyl glutarate (DSG), after which fixed chromatin was digested with the sequence-independent endonuclease DNase I. Chromatin ends were repaired and ligated to a biotinylated bridge adapter, before proximity ligation of adapter containing ends. After ligation, crosslinks were reversed, and the DNA was purified from proteins. Purified DNA was treated to remove biotin that was not internal to ligated fragments. Dual indexed primers supplied in the Dovetail Dual Index Primer Set #1 for Illumina Module were used for indexing. Shallow sequencing was performed for each Omni-C library as described in the QC analysis pipeline (https://omni-c.readthedocs.io/en/latest/) before proceeding to target enrichment.

#### pCHi-C target enrichment

pCHi-C libraries were generated using the Dovetail® Human Pan Promoter Enrichment Kit according to the manufacturer’s instructions (Dovetail Genomics, 25013). The Dovetail Genomics Custom Panel covers 84,643 promoters associated with 27,375 coding and non-coding genes. Since the total panel size is between 10 and 50 Mb, eight PCR cycles were used for the post-hybridisation PCR amplification.

#### pCHi-C next generation sequencing (NGS), mapping and filtering

pCHi-C libraries were sequenced on an Illumina HiSeq X Ten System (Illumina) generating 150 bp paired-end (PE) reads. Sequences were output in fastq format before mapping against the human reference genome (GRCh38) generating between 117-470 million di-tags (Supplementary Table 1). Filtering to remove experimental artefacts/duplicates was carried out using the publicly available Dovetail Omni-C pipeline (summary statistics including the proportion of PCR duplicates, off target pairs and distance distribution of *cis* di-tags are shown in Supplementary Table 1).

The Dovetail Pan Promoter Enrichment Panel targets >98% of human promoter regions (∼84,000 promoter sequences) and, for the purposes of analysis, allocates these to 39,825 genomic (“promoter”) bins that capture the entire promoter sequence and negate promoter sequences split across bins. The majority of these promoter bins capture promoter(s) at a single gene but there is a subset (n=2,905) where the proximity of gene promoters is such that a single promoter bin captures multiple promoters and there is a subset of genes (n=9,104) for which there is more than one promoter bin. For the purpose of downstream analysis multiple promoter bins for a single gene have been numbered in ascending order (“P#”) according to their 5’ to 3’ orientation. After filtering for alignment quality and PCR duplicates, off target di-tags (defined as di-tags where neither end mapped to a promoter bin) were removed leaving 20 to 88 million unique valid di-tags per sample for downstream analyses.

#### Assessing reproducibility

To test replicate reproducibility, we followed our previously published approach^14,33^ of examining distance stratified numbers of read counts between replicate pairs. Replicate concordance quantified as Spearman’s ρ varied from 0.76-0.94 at short distances (0-10kb), 0.22-0.42 at the distances over which we would expect to detect long range interactions (100kb-1Mb) and 0.08-0.20 at distances where the data will be sparse (1Mb-10Mb) (Supplementary Figures 5-9). There was no evidence that concordance was greater in technical replicates than in biological replicates, with greater concordance between high quality libraries than in libraries with less consistent quality as assessed by percentage of PCR duplicates, percentage of off target di-tags and percentage of short range (≤1kb) interactions (Supplementary Table 1).

#### Interaction peak calling

Replicate counts for each cell type were combined for interaction peak calling with CHiCANE^14^ (R package v0.1.9) using the default parameters. Bait-to-bait interaction peaks were excluded, and only significant interaction peaks (q-value ≤ 0.1) were considered for downstream analysis. All non-significant and bait-to-bait interactions are retained in raw data contact heatmaps.

#### H3K27ac CUT&Tag data generation

Cells (HMLE, MCF10A, T-47D, HMF and GS2) were prepared for CUT&Tag following the protocol from Active Motif (https://www.activemotif.com/catalog/1320/cut-tag-service). Briefly, adherent cells were washed in 1X PBS and collected in 1X Enzyme Free Cell Dissociation Solution Hank’s Based (Sigma) using a cell scraper at room temperature. Cells were counted using a Countess II Automated Cell Counter (Thermo Fisher), pelleted at 500g for 5 minutes at 4°C, and resuspended in ice-cold preservation solution (50% FBS/40% growth media/10% DMSO (Sigma)) at a concentration of 400 cells/µl. 500µl (200,000 cells) was transferred to a 1.5ml tube on ice and cells were gradually frozen to -70°C in a styrofoam container. Cells were shipped on dry ice to Active Motif for H3K27ac CUT&Tag data generation. Briefly, cells were incubated overnight with concanavalin A beads and 1µl of the primary anti-H3K27ac antibody per reaction (#39135, Active Motif). After incubation with the secondary anti-rabbit antibody (1:100), cells were washed and tagmentation was performed at 37°C using protein-A-Tn5. Tagmentation was halted by the addition of EDTA, SDS and proteinase K at 55°C, after which DNA extraction and ethanol purification was performed, followed by PCR amplification and barcoding (see Active Motif CUT&Tag kit, #53160 for recommended conditions and indexes). Following SPRI bead cleanup (Beckman Coulter), the resulting DNA libraries were quantified and sequenced on an Illumina NextSeq 550 (8 million reads, 38 bp PE reads).

#### H3K27ac CUT&Tag data analysis

Reads were aligned using the BWA algorithm (mem mode; default settings)^38^. Duplicate reads were removed and only reads that mapped uniquely (mapping quality ≥ 1) and as matched pairs were used for further analysis. Peaks were identified using the MACS 3.0.0 algorithm at a cutoff FDR corrected P-value of 0.05, without control file, and with the –nomodel option. Peaks that were on the ENCODE blacklist of known false ChIP-seq peaks were removed. Signal maps and peak locations were used as input data to Active Motif’s proprietary analysis program, which creates output containing information on sample comparison, peak locations and gene annotations. Consensus peaks per cell type were defined as the maximum coordinates of peaks present by intersect in both replicates.

#### H3K4me3 data

H3K4me3 ChIP-seq data generated in human mammary epithelial cells and human mammary fibroblasts were downloaded from ENCODE 4, hg38: https://www.encodeproject.org/files/ENCFF898NGW/ (H3K4me3 ChIP-seq on human HMEC) and https://www.encodeproject.org/files/ENCFF073WBH/ (H3K4me3 ChIP-seq on human HMF). Data were downloaded as narrowPeaks, without further processing and were aligned with pCHi-C bait and target bins.

#### RNA-seq generation

RNA extraction from cells was performed according to the manufacturer’s instructions using RNeasy Mini Kit (Qiagen). RNA was quantified using Qubit RNA BR Assay Kit (Invitrogen) and quality was assessed using TapeStation High Sensitivity RNA ScreenTape (Agilent). At the ICR Genomics Facility, intact RNA was processed using mRNA (polyA) selection, treated with TURBO DNase (Invitrogen) and the library prepared for sequencing using NEBNext Ultra II Directional RNA Library Prep Kit for Illumina (NEB). Isoform specific RNA-seq data generation from the CRISPRi samples was performed by Genewiz (Azenta Life Sciences) (standard RNA-seq with polyA selection, 150M reads per sample).

#### Gene expression analysis by RNA-seq

RNA-seq profiling generated 28.1-37.7 million PE reads for the primary cell datasets (HMLE and HMF), and 135.8-215.9 million PE reads for the CRISPRi RNA datasets. Fastq reads were trimmed using Trim Galore (v0.6.6). FastQC, FastQ Screen^39^ and MultiQC (v1.9)^40^ were run to determine library quality. PE reads (150bp) were aligned to the human reference genome GRCh38 using STAR 2.7.6a^41^ with --quantMode GeneCounts and --twopassMode Basic alignment settings with annotation files downloaded from GENCODE (v22) in GTF file format. Results were further annotated using ENSEMBL gene annotations with the R package org.Hs.eg.db (v3.10.0) in the R statistical programming environment (v3.6.0). Differential gene expression analysis was performed using edgeR (v3.28.1). Differential exon usage analysis was performed using DEXSeq pipeline (R package v1.32.0).

#### NEBNext Enzymatic Methyl-seq (EmSeq) data generation

DNA extraction from cells (HMLE and HMF) was performed using QIAamp DNA Mini Kit (Qiagen) and quantified using Qubit dsDNA Quantification Assay Kit (Invitrogen) and Qubit 3 Fluorometer (Invitrogen). NEBNext EmSeq libraries were prepared from 200ng of DNA with 0.001ng of CpG methylated pUC19 and 0.02ng of unmethylated λ phage DNA (NEB) spiked in to measure enzymatic conversion efficiency. Samples were fragmented using a Covaris E220 focused-ultrasonicator to an average size of 400bp. Conversion was performed using the NEBNext EmSeq Conversion Module (NEB) and was >99.5% in all samples, as measured by the internal controls. Library preparation was performed following the Protocol for use with Large Insert Libraries (470-520bp) from NEBNext EmSeq Kit (NEB). Library quality was monitored using TapeStation 4150 System (Agilent). Whole genome sequencing was carried out by the ICR Genomics Facility on the Illumina NovaSeq 6000 System with 30X coverage.

#### NEBNext EmSeq data analysis

Read quality of each EmSeq whole genome sequencing fastq file was assessed with multiQC (v1.9). EmSeq forward and reverse reads were aligned to a hybrid reference genome containing human (GRCh38), pUC19 and λ phage (J02459.1) using bwa-meth (v0.2.2). Subsequent BAM files were sorted and indexed with SAMtools (v1.11). Duplicate reads were marked and removed with MarkDuplicates (gatk v4.1.9.0). CollectRrbsMetrics (gatk v4.1.9.0) was used to extract summary and metrics tables including coverage distribution and EmSeq base conversion rate. Methylation bias identification, plotting and correction was performed with MethylDackel (v0.5.3) (https://github.com/dpryan79/MethylDackel). A bespoke bash script was used to extract the appropriate read correction parameters from MethylDackel mbias output. CpG conversion rates over pUC19 and λ phage J02459.1 were used as further QC of library quality.

CpG island annotation BED files required for annotation of differential methylated regions were downloaded from UCSC table browser (https://genome.ucsc.edu/cgi-bin/hgTables) as per instructions on https://github.com/al2na/methylKit. Promoter bins are as previously described in **pCHi-C next generation sequencing (NGS), mapping and filtering**. Promoter bin and CpG-specific QC and differential methylation analysis was performed using methylKit (v1.18.0)^42^. All sites with less than 5X coverage were discarded along with bases with coverage higher than the 99.9^th^ percentile of coverage in each sample. BedGraph files were generated by methylKit bedGraph() for viewing normalised sample percent methylation and differential methylation sites in IGV (v2.11.1) or UCSC. All statistical analysis was performed in R (v4.1.0).

#### Genotyping

The genotype of CCVs (Supplementary Table 4) was determined by PCR amplification followed by Sanger sequencing. Genomic DNA was extracted from MCF10A, T-47D, GS2, 1989 HMF and 3002 HMF cells in culture using Qiagen QIAamp DNA Blood Mini Kit (Qiagen) according to the manufacturer’s instructions and used as template in a hi-fidelity PCR reaction. Primers (Supplementary Table 9) were used to amplify a DNA fragment, with 1X Q5 Reaction Buffer, 200µM dNTPs (Invitrogen), 0.5µM each forward and reverse primer (Sigma-Aldrich), 0.02U/µl Q5 High-Fidelity DNA Polymerase (New England Biolabs), 50ng gDNA, and thermocycle conditions: 98°C, 30sec; followed by 35 cycles of: 98°C, 5sec; T_an_°C (Supplementary Table 9), 20sec; 72°C, 20sec; followed by 72°C, 2min; with a final hold at 4°C. The PCR product was run on a 2% agarose/1XTAE gel and purified using QIAquick Gel Extraction Kit according to the manufacturer’s instructions. 5µl purified DNA (10-50ng/µl) was premixed with 5µl primer (5µM) and sequenced in both forward and reverse direction at Genewiz (Azenta Life Sciences).

#### Western blotting

Cells were washed in PBS and whole cell extracts were prepared by lysing cells with lysis buffer (150mM NaCl, 50mM Tris pH 8.0, 1% Triton X-100, PhosStop and Complete Mini EDTA free protease inhibitors (Roche)) on ice for 5 min. Lysed cell suspension was collected using a cell scraper and agitated for 30 mins at 4°C. Following centrifugation at 13,000rpm for 25 mins at 4°C, the whole cell lysate supernatant was aliquoted and stored at -70°C. Protein yield was quantified by Bio-Rad Protein Assay Dye Reagent Concentrate (Bio-Rad) with BSA (NEB) as standard.

Whole cell lysates were subjected to SDS-PAGE on a 4-12% Bis Tris gel with MOPS buffer (Invitrogen) and transferred to a nitrocellulose membrane (0.45 µm pore size, Invitrogen). The membrane was blocked with 5% non-fat milk in TBST (1X TBS, 0.05% Tween 20) at room temperature overnight with shaking. Membranes were probed with antibodies against FILIP1L (HPA043133, Sigma) diluted 1:1,000; or vinculin (as loading control; sc-73614, Santa Cruz Biotechnology) diluted 1:5,000; in 5% non-fat milk in TBST for 2 hours at room temperature. After washing 3 times with TBST, the blots were incubated with anti-rabbit IgG secondary antibody (A0545, Sigma) or anti-mouse IgG secondary antibody (A9044, Sigma) conjugated to horseradish peroxidase diluted 1:10,000 in TBST for 1 hour at room temperature. The blots were then washed 4 times with TBST and developed with ECL Detection reagent (Cytiva).

#### JESS Simple Western™ capillary-based immunoassay

Sample separation and detection was performed on the JESS Simple Western™ instrument (ProteinSimple^®^, Bio-Techne) according to the manufacturer’s instructions using Fluorescence Separation Module SM-FL004, containing the 12-230 kDa 25-Capillary Cartridge, and Chemiluminescence Detection Module DM-001, containing Goat Anti-Rabbit Secondary HRP antibody (#042-206). Briefly, whole cell lysates were diluted to 0.13 µg/µl in 0.1X Sample Buffer, mixed 4:1 with 5X Fluorescent Master Mix (EZ standard pack 1; #PS-ST01EZ, ProteinSimple^®^) and 3 µl added per well of the microplate. The polyclonal rabbit anti-FILIP1L antibody (#HPA043133, Sigma) and the polyclonal rabbit anti-beta Tubulin antibody (#ab6046, Abcam) were multiplexed at 1:200 and 1:2000 dilution respectively in Antibody Diluent 2 and 10 µl added per well.

JESS Simple Western™ data were analysed using Compass for Simple Western software. Peaks were automatically detected. Images from the High Dynamic Range 4.0 were used for the analysis. For quantification, FILIP1L and beta tubulin peak areas were log2 transformed. Log2 fold changes in FILIP1L, adjusted for beta tubulin, for each sgRNA compared to no sgRNA were calculated using analysis of variance.

#### FILIP1L cDNA transfection

T-47D cells were transfected using Lipofectamine 3000 (Invitrogen) according to the manufacturer’s instructions. Briefly, cells were seeded in a T75 flask to be 80% confluent at transfection with (i) water (mock), (ii) 15µg pEGFP-C1 (Clontech) (transfection control), (iii) 15µg FILIP1L NM_001282793 (ENST00000487087) expression plasmid (SC337274, Origene), (iv) 15µg FILIP1L NM_014890 (ENST00000383694) expression plasmid (SC114738, Origene). Whole cell lysates were prepared 48 hours post transfection.

#### CRISPR-based perturbation

Single guide RNAs (sgRNAs) were designed against target sequences using the online design tool CHOPCHOP (http://chopchop.cbu.uib.no). Target sequences were selected based on their proximity to variants or regions of interest and specificity scores (Supplementary Table 6). Cloning was performed as described by Ran *et al*^43^. Briefly, guides were synthesised (Merck) as two complementary oligonucleotides with 5’-CACC and 5’-AAAC (top and bottom strand oligonucleotides, respectively) overhangs to facilitate cloning; a G nucleotide was substituted at the first base of the target sequence if not already present to improve transcription from the U6 promoter. 1µl of each oligo (100µM) was phosphorylated and annealed with 5U T4 Polynucleotide Kinase (NEB) and 1X T4 ligation buffer (NEB) in a 10µl reaction. Annealing was carried out in a PCR machine with programme: 37°C, 30min; followed by 95°C, 5min; followed by ramp down to 25°C at 0.1°C/sec; with a final hold at 4°C. Cloning of annealed oligos was performed following linearisation of 2µg guide expression vector pKLV2-U6gRNA5(BbsI)-PGKpuro2AZsG-W (#67975, Addgene;^44^) with 30U BbsI (NEB) and 1X NEBuffer r2.1 (NEB) in a 50µl reaction. In a 10µl reaction, 20ng of the gel purified linear vector, 20U T4 ligase (NEB), 1X ligase buffer (NEB), 2µl annealed oligos (diluted to 7.1fmol/µl in EB Buffer (Qiagen)) was incubated at 16°C overnight and 5µl of the ligation mixture was transformed into 50µl chemically competent bacterial cells. Cloning was validated by Sanger sequencing (Eurofins Genomics).

For lentiviral packaging, pKLV2-U6gRNA5(BbsI)-PGKpuro2AZsG-W containing cloned guides, or Lenti-dCas9-KRAB-blast (#89567, Addgene), or pGH125_dCas9-Blast (#85417, Addgene) (for generation of cells stably expressing dCas9-KRAB or dCas9, respectively) were packaged into lentiviral particles in 293T cells. Briefly, 1.2x10^6^ cells per well were seeded in a 6-well plate and cultured overnight. Two reaction mixes were prepared: (1) 4µl Lipofectamine 3000 (Thermo Fisher Scientific), 125µl OptiMEM (Thermo Fisher Scientific); (2) 500ng plasmid to be packaged, 500ng psPAX2 (#12260, Addgene), 150ng pMD2.G (#12259, Addgene), 125µl OptiMEM, 2.5µl P3000 reagent (Thermo Fisher Scientific). These were combined and incubated at room temperature for 20 minutes. The mixture was added dropwise to the cells and returned to culture. After 24 hours the media was replaced. After an additional 48 hours the media containing lentivirus particles was collected, stored at 4°C, and replaced with fresh media. After a further 24 hours the media containing lentivirus particles was collected, pooled with the first aliquot and centrifuged at 1600g, 10 minutes, 4°C to pellet cellular debris. The supernatant was filtered using Millex-HV 0.45 μM PVDF filters (Millipore). Before storage, lentiviral supernatant containing sgRNAs was concentrated by combining 3 volumes of clarified supernatant with 1 volume of Lenti-X Concentrator (Clontech). They were mixed by gentle inversion, incubated at 4°C for 30 minutes, and centrifuged at 1500g for 45 minutes at 4°C. After centrifugation, an off-white pellet was resuspended in 1/10 of the original volume using PBS and aliquoted for storage at -70°C.

The GS2 CRISPRi modified cell line was derived from parental GS2 cell lines. Stable expression of dCas9-KRAB (catalytically dead Cas9 fused to the KRAB transcriptional repressor) was introduced by transduction with lentiviral particles prepared as described above^45^. Briefly, cells were seeded in a T75 flask, so they were 50% confluent on the day of transduction. 500µl lentiviral supernatant was added to the cells and incubated overnight. After 24 hours, the media was replaced with normal media and 48 hours after transduction successfully transduced cells were selected for by treatment with 10µg/ml blasticidin (InvivoGen). Expression of dCas9-KRAB was confirmed by Western blot with an antibody against dCas9 ((7A9-3A3) N-Terminus-BSA Free, NBP2-36440, Novus Biologicals) (Supplementary Figure 10). dCas9-KRAB expressing cells were seeded into 12-well plates at a density of 100,000 cells per well. After 24 hours, 100µl sgRNA lentiviral particles was added. After 96 hours, successfully transduced cells expressing ZsGreen (visualised by fluorescent microscope) were selected for by replacement with fresh media containing 1µg/ml puromycin (InvivoGen).

#### Figure generation

All figures were generated in the R v3.6.0 environment using custom scripts. Euler plots were generated using eulerr v6.1.0. Loci interaction figures and loci CpG methylation figures were generated using Gviz v1.30.3, AnnotationDbi_ v.48.0, GenomicInteractions v1.20.3, GenomicFeatures v1.38.2, RColorBrewer v1.1-2, EnsDb.Hsapiens v86_2.99.0, GenomicRanges_ v.38.0, rtracklayer v1.46.0 and IRanges v2.20.0. Custom post-hoc editing of aesthetics was performed in Inkscape v1.2.

## Declarations

### Ethics approval and consent to participate

All primary human breast tissue was obtained from the Breast Cancer Now Tissue Bank (REC 21/EE/0072).

## Consent for publication

Not applicable.

## Availability of data and materials

Raw capture pCHi-C (HMLE x2, HMF x2, MCF10A x3, T-47D x2, GS2 x2), H3K27ac CUT&Tag (MCF10A x2, T-47D x2, GS2 x2, HMLE x2, HMF x2), whole genome NEBNext EmSeq (HMLE x2, HMF x2), bulk RNA-seq (HMLE x5, HMF x4,) and CRISPRi-modified RNA-seq (GS2 x21) generated in this study are deposited in the Sequence Read Archive under the submission number PRJNA1235642. Processed pCHi-C interaction peaks (HMLE x1, HMF x1, MCF10A x1, T-47D x1, GS2 x1) generated in this study are deposited in the Gene Expression Omnibus under the submission number GSE291943. Processed pCHi-C interaction peaks can be uploaded to the WashU Epigenome Browser (https://epigenomegateway.wustl.edu/) in the “Tracks” function by selecting “Local text tracks” and then the “Long range format by CHiCANE” option from the “Choose text file type” drop down menu. CHiCANE interaction peak files must be unzipped prior to WashU upload; the WashU browser will accept zipped files, but they will appear to contain no interactions. Processed H3K27ac CUT&Tag consensus peaks (HMLE x1, HMF x1, MCF10A x1, T-47D x1, GS2 x1) generated in this study are deposited in the Gene Expression Omnibus under the submission number GSE291944. The Genotype-Tissue Expression (GTEx) Project was supported by the Common Fund (https://commonfund.nih.gov/GTEx) of the Office of the Director of the National Institutes of Health and by NCI, NHGRI, NHLBI, NIDA, NIMH, and NINDS. The data used for the analyses described in this manuscript were obtained from the GTEx Portal on 01/25/2024.

## Author Contributions

Conceptualization: OF, SH, NJ. Methodology: OF, SH, NJ, HK. Formal analysis: OF, HK, AG, SH, JA. Investigation: AZ, NJ, KT, SRC, VF, SS, KM, JSB, CJL, SJP, JLJ, JG, IG. Data curation: SH, HK. Writing – original draft: AZ, HK, OF, SH, NJ. Writing – review and editing: all authors. All authors read and approved the final manuscript.

## Acknowledgements

This work was supported by Programme Grants from Breast Cancer Now as part of Programme Funding to the Breast Cancer Now Toby Robins Research Centre and by an ICR studentship to OF and SH. We thank the Breast Cancer Now Toby Robins Research Centre Research Bioinformatics Group for Bioinformatics support and thank Breast Cancer Now, working in partnership with Walk the Walk, for supporting the work of this team. We acknowledge National Health Service funding to the NIHR Royal Marsden Biomedical Research Centre.

We thank Professor Clare Isacke for providing GS2 mammary fibroblasts and Rebecca Orha and Marjan Iravani for advice and assistance with GS2 cell culture. The authors wish to acknowledge the role of the Breast Cancer Now Tissue Bank in collecting and making available the samples used in the generation of this publication, and all the patients who donated the samples. We thank Dovetail Genomics (now part of Cantata Bio LLC) for support with pCHi-C library sequencing and the ICR Genomics Facility for support with pCHi-C library, NEBNext Enzymatic Methyl-seq (EmSeq) library and RNA-seq library sequencing.

For the purpose of Open Access, the author has applied a CC BY public copyright licence to any Author Accepted Manuscript (AAM) version arising from this submission.

pKLV2-U6gRNA5(BbsI)-PGKpuro2AZsG-W was a gift from Kosuke Yusa (Addgene plasmid # 67975; http://n2t.net/addgene:67975; RRID:Addgene_67975). Lenti-dCas9-KRAB-blast was a gift from Gary Hon (Addgene plasmid # 89567; http://n2t.net/addgene:89567; RRID:Addgene_89567). pGH125_dCas9-Blast was a gift from Michael Bassik (Addgene plasmid # 85417; http://n2t.net/addgene:85417; RRID:Addgene_85417). psPAX2 (Addgene plasmid # 12260; http://n2t.net/addgene:12260; RRID:Addgene_12260) and pMD2.G (Addgene plasmid # 12259; http://n2t.net/addgene:12259; RRID:Addgene_12259) were gifts from Didier Trono.

